# The Polygenic and Monogenic Basis of Blood Traits and Diseases

**DOI:** 10.1101/2020.02.02.20020065

**Authors:** Dragana Vuckovic, Erik L. Bao, Parsa Akbari, Caleb A. Lareau, Abdou Mousas, Tao Jiang, Ming-Huei Chen, Laura M. Raffield, Manuel Tardaguila, Jennifer E. Huffman, Scott C. Ritchie, Karyn Megy, Hannes Ponstingl, Christopher J. Penkett, Patrick K. Albers, Emilie M. Wigdor, Saori Sakaue, Arden Moscati, Regina Manansala, Ken Sin Lo, Huijun Qian, Masato Akiyama, Traci M. Bartz, Yoav Ben-Shlomo, Andrew Beswick, Jette Bork-Jensen, Erwin P. Bottinger, Jennifer A. Brody, Frank J.A. van Rooij, Kumaraswamy N. Chitrala, Kelly Cho, Hélène Choquet, Adolfo Correa, John Danesh, Emanuele Di Angelantonio, Niki Dimou, Jingzhong Ding, Paul Elliott, Tõnu Esko, Michele K. Evans, Stephan B. Felix, James S. Floyd, Linda Broer, Niels Grarup, Michael H. Guo, Andreas Greinacher, Jeff Haessler, Torben Hansen, Joanna M. M. Howson, Wei Huang, Eric Jorgenson, Tim Kacprowski, Mika Kähönen, Yoichiro Kamatani, Masahiro Kanai, Savita Karthikeyan, Fotis Koskeridis, Leslie A. Lange, Terho Lehtimäki, Allan Linneberg, Yongmei Liu, Leo-Pekka Lyytikäinen, Ani Manichaikul, Koichi Matsuda, Karen L. Mohlke, Nina Mononen, Yoshinori Murakami, Girish N. Nadkarni, Kjell Nikus, Nathan Pankratz, Oluf Pedersen, Michael Preuss, Bruce M. Psaty, Olli T. Raitakari, Stephen S. Rich, Benjamin A.T. Rodriguez, Jonathan D. Rosen, Jerome I. Rotter, Petra Schubert, Cassandra N. Spracklen, Praveen Surendran, Hua Tang, Jean-Claude Tardif, Mohsen Ghanbari, Uwe Völker, Henry Völzke, Nicholas A. Watkins, Stefan Weiss, VA Million Veteran Program, Na Cai, Kousik Kundu, Stephen B. Watt, Klaudia Walter, Alan B. Zonderman, Peter W.F. Wilson, Yun Li, Ruth J.F. Loos, Julian Knight, Michel Georges, Oliver Stegle, Evangelos Evangelou, Yukinori Okada, David J. Roberts, Michael Inouye, Andrew D. Johnson, Paul L. Auer, William J. Astle, Alexander P. Reiner, Adam S. Butterworth, Willem H. Ouwehand, Guillaume Lettre, Vijay G. Sankaran, Nicole Soranzo

## Abstract

Blood cells play essential roles in human health, underpinning physiological processes such as immunity, oxygen transport, and clotting, which when perturbed cause a significant health burden. Here we integrate data from UK Biobank and a large-scale international collaborative effort, including 563,946 European ancestry participants, and discover 5,106 new genetic variants independently associated with 29 blood cell phenotypes covering the full allele frequency spectrum of variation impacting hematopoiesis. We holistically characterize the genetic architecture of hematopoiesis, assess the relevance of the omnigenic model to blood cell phenotypes, delineate relevant hematopoietic cell states influenced by regulatory genetic variants and gene networks, identify novel splice-altering variants mediating the associations, and assess the polygenic prediction potential for blood cell traits and clinical disorders at the interface of complex and Mendelian genetics. These results show the power of large-scale blood cell GWAS to interrogate clinically meaningful variants across the full allelic spectrum of human variation.

## Introduction

A major aspiration in the field of human genetics is to understand how human genetic variation impacts complex traits and diseases. Recent genome-wide association studies (GWAS) have identified thousands of genetic variants associated with complex phenotypes and provided insights into their genetic architecture. This has led to the recognition that complex trait heritability is polygenic, resulting from the cumulative effects of many genetic loci, each of modest effect size, located throughout the genome (Timpson et al., 2018; Visscher et al., 2017).

Hematopoiesis is a valuable example for studying complex trait genetic architecture, since blood cell phenotypes are commonly measured in large population-based studies and the production of blood cells (red blood cells, white blood cells, and platelets) is a highly regulated, hierarchical process that is largely intrinsic to hematopoietic progenitor cells and intermediate cell states, which can be readily isolated (Bao et al., 2019; Tardaguila and Soranzo, 2019). While there have been advances in understanding genetic loci associated with blood cell production, the full allelic spectrum of human genetic variation impacting this process remains incompletely understood. Population-scale studies of hundreds of thousands, and eventually millions, of participants offer a unique opportunity to catalogue the spectrum of human variation impacting hematopoiesis.

Most genetic variants contributing to complex trait heritability are noncoding variants located in genomic regulatory regions within relevant cell types. The availability of epigenomic and transcriptomic profiles for hematopoietic stem/ progenitor and lineage-committed cells enable mechanistic dissection of the role that different classes of genes have in the process of hematopoiesis. Prior studies of blood cell traits have suggested that master transcription factors may be impacted by genetic variation (Ulirsch et al., 2019) and it is likely that further studies may uncover additional roles for, and variation of, key regulators affecting these processes. Another priority is to advance understanding of network connectivity between trait-associated genes and variants, and this understanding can be informed by theoretical models. Recently, an ‘omnigenic’ model has been proposed in which two types of genes (‘core’ vs ‘peripheral’) differentially contribute to complex trait heritability (Boyle et al., 2017; Liu et al., 2019). However, the extent to which the omnigenic model is applicable to various complex traits and diseases remains unclear and questions have emerged regarding the broad applicability of this model (Wray et al., 2018).

Finally, although rare variants with large effects generally do not individually contribute substantially to overall complex trait variance, they can often highlight important biologic mechanisms. The emergence of large population-based data sets with dense genotyping and high-quality imputation of rare genetic variants can enable comprehensive capture of the full spectrum of genetic variation contributing to complex traits. For example, some of the same rare variants of large effect may contribute to rare hematologic disorders, many of which are characterized by variable penetrance or expressivity. Polygenic contributions of many variants with small effects can yield disease risk odds ratios comparable to or larger than that of known monogenic variants (Oetjens et al., 2019). Therefore, large population-based data sets can also help to re-classify the pathogenicity and penetrance of disease-causing variants (including variants of uncertain clinical significance, VUS), as well as understand the contribution of polygenic variation to risk of blood diseases or as modifiers of rare variants that contribute to Mendelian blood disorders.

## Results

### Genetic variants associated with blood count phenotypes

We leveraged the power of the UK Biobank to perform a genome-wide discovery analysis in N=408,112 participants of European ancestry, investigating 29 blood cell phenotypes (**Supplementary Table 1**). In parallel, we also performed tests for genetic associations with a subset of 15 phenotypes available in an additional 159,973 European ancestry participants from the Blood Cell Consortium (BCX; **Figure 1A, Supplementary Table 2**). A separate analysis of non-European participants is reported in a companion paper (Chen et al., 2020). Overall, this discovery effort identified 16,643 autosomal and 257 X-linked conditionally-independent (**Methods**) trait-variant associations (**Figure 1B, Supplementary Table 3 and 4**). The 16,900 associations were assigned to 7,122 genomic loci (5,106 not described before) using an LD clumping approach (Astle et al., 2016). Each represented by a unique tag variant (between-tag pairwise LD r^2^≤ 0.8), and for simplicity, throughout the paper we use the term ‘sentinel variant’ to refer to either a clump tag variant or a trait specific conditionally independent signal. Overall, we nearly tripled the number of blood cell loci reported prior to this study (Astle et al., 2016). We assessed replication rates across three exemplar phenotypes (platelet counts, lymphocyte counts and red cell counts) for 210 variants on chromosome 1 in the Million Veterans Program (MVP, N=271,280) and found that all of them had concordant effect size estimates (Pearson’s R^2^=0.94) and 196 (93%) variants replicated at a nominal significance threshold (p<0.05), with the non-replicating ones lacking power due to small sample size (**Supplementary Figure 1A**). Using a Bayesian method that accounts for multiple independent signals (Benner et al.) (**Methods, Figure 1C**), we fine-mapped 3,100 (19% of 16,643 autosomal) associations to a single putative causative variant (>95% posterior probability [PP_FM_])(**Supplementary Table 5**), and more than half of the associated signals (n=9,149, 55%) to fewer than 10 variants (**Figure 1D**). As expected, rare signals are more likely to be fine-mapped to smaller credible sets (**Figure 1D**). We annotated variants to genes using the variant effect predictor (VEP) worst consequence annotation (McLaren et al., 2016). Among the sentinel variants, 8,866 (83% of unique sentinels) were annotated to a gene using this approach, of which 69% were intronic, 24% were in regulatory regions, and 7% were in protein-coding regions (5.5% non-synonymous and 1.5% synonymous; **Figure 1E**). The credible set size distribution (number of variants per credible set) was consistent across traits (**Figure 1F**). Furthermore, we used colocalization analyses (Giambartolomei et al., 2018) to systematically compare each association signal to cis-eQTLs derived from 6 different blood cell types relevant to each trait (platelets n=424; CD19^+^B-cells, CD8^+^T-cells, CD4^+^T-cells and CD15 neutrophils n=300; CD14 monocytes n=1,490). The concordance of genes assigned using VEP to eGenes at 667 colocalizing eQTLs was 65% (Kreuzhuber, 2019)(**Supplementary Figure 1B**).

**Figure 1:**
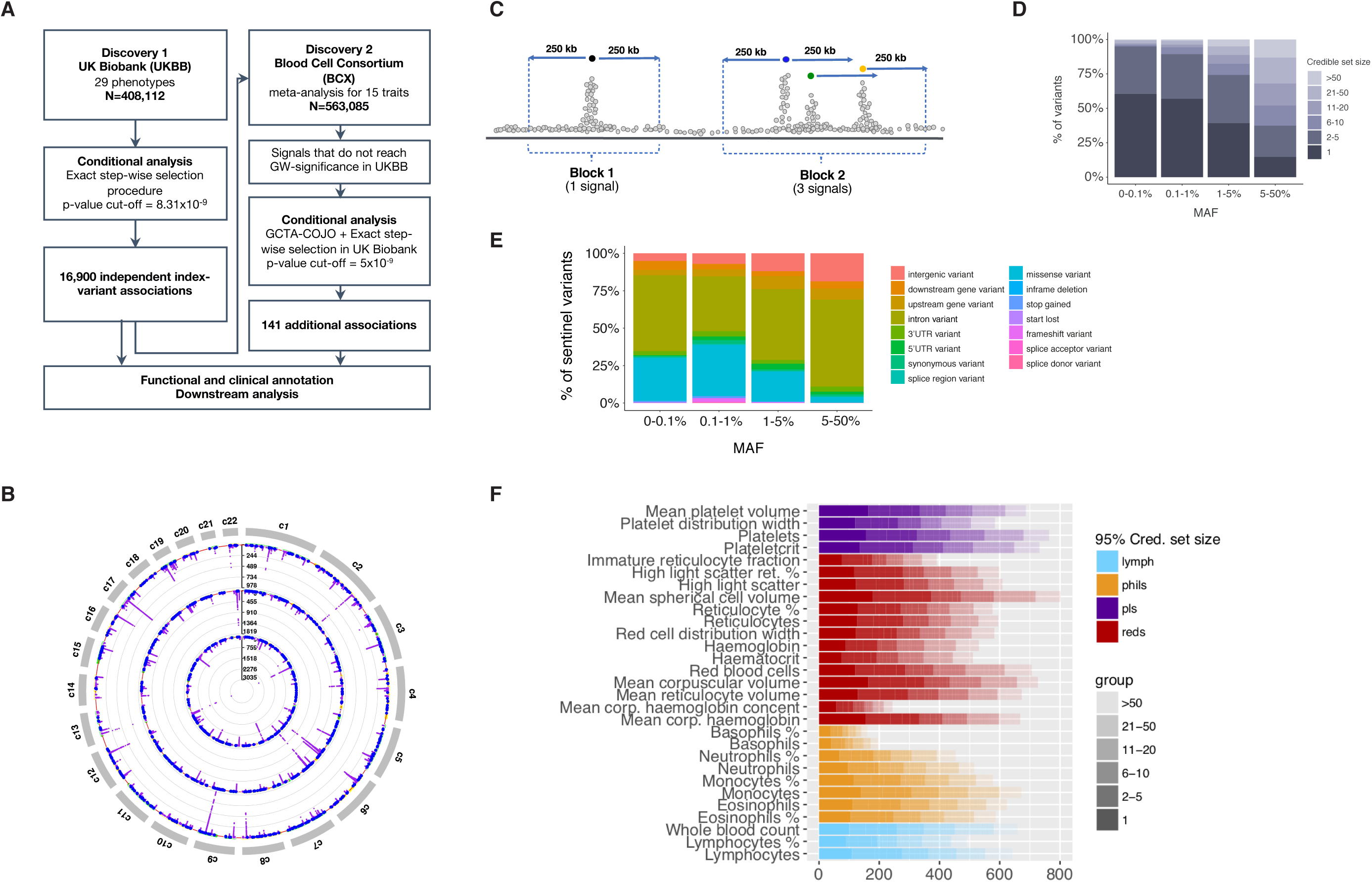
GWAS study design and results. **A**, study design; **B**, Circular Manhattan plot for the meta-analysis results (Discovery 1+2). 15 blood cell traits are grouped into White Blood Cells (WBC), Red Blood Cell (RBC) and Platelets (PLT). For each of the three groups, minimum p-value from all traits in that group is presented if it is <10^−7^. The red line represents the significance threshold of 5×10^9^. Purple points have p<5×10^−9^ and blue points are novel associations. Yellow and green points have intermediate 10-7<p<5×10^−9^; **C**, Illustrative cartoon for fine-mapping strategy showing how the FM blocks and the relevant number of causative signals were defined; **D**, Distribution of fine-mapping results by minor allele frequency (MAF); **E**, Distribution of fine-mapping results by sentinel variant annotation and MAF; **F**, Fine-mapping 95% credible set size distribution for each conditionally independent signal, across all traits: different colors indicate different cell type groups.

### Genetic architecture and network connectivity of blood cell traits

Hematopoiesis is a finely-tuned and highly coordinated process involving hundreds of co-regulated genes, and it is likely that a subset of the genetic variants associated with peripheral blood cell counts and indices acts upon master regulators of this process. We therefore sought to evaluate the extent to which genes assigned to our GWAS signals were represented in a whole blood gene coexpression network (Nath et al., 2017), which includes 7,509 connected genes (**Methods**). When genes were assigned to GWAS signals using VEP worst consequence annotation as detailed above, one quarter of the network genes (25%, n=1,874 genes) could be linked to a sentinel variant. Expanding the VEP annotation to include any gene consequence yielded an additional 2.5% (n=196) genes (**Figure 2A)**. When considering all genes included in a 250 kb fine-mapping block centered on the sentinel variant, 78% of network genes could be assigned to a GWAS locus, representing a significant enrichment compared to random sets of genes (p<10^−4^by permutation test, **Figure 2B-C, Supplementary Table 6**). Overall, 88% of sentinel variants were close (<250 kb) to a network gene, suggesting that knowledge of the network structure can aid the assignment of likely effector genes to non-coding associations.

**Figure 2.**
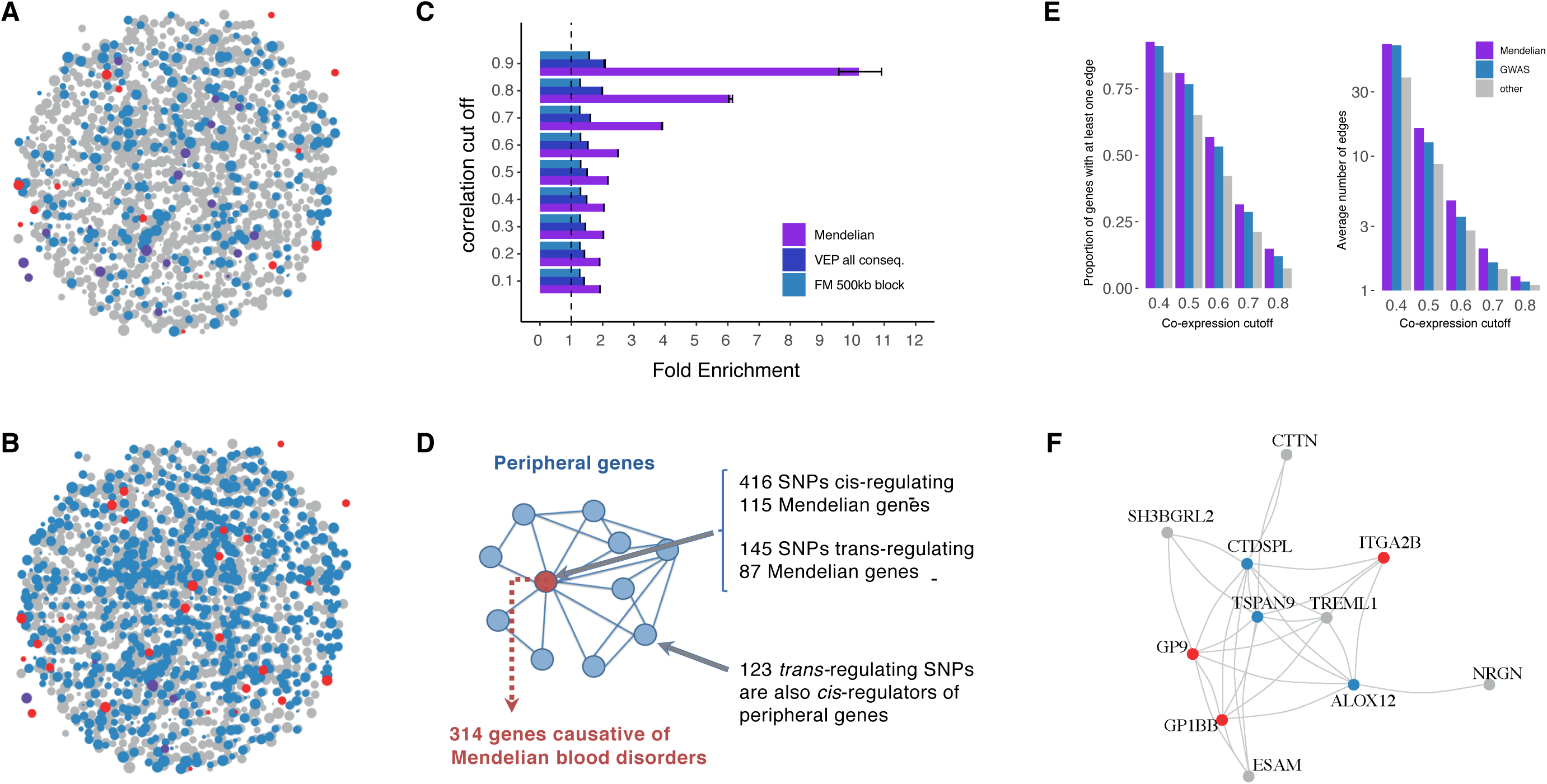
Network connectivity. **A-B**, Co-expression network in whole blood. Genes shown have at least one edge with another gene and co-expression correlation > 0.7. Edges are omitted for clarity, instead the node size summarizes the number and strength of co-expression links. Blue dots represent genes detected by GWAS, violet dots represent Mendelian genes and red dots represent the intersection. Grey dots are genes in the co-expression network that do not belong to any of the previous categories. GWAS genes are defined by two different variant annotation approaches: VEP all consequences (**A**) and 500kb fine-mapping windows (**B**); **C**, Enrichment of sets of genes in the co-expression network, compared to permutations of random sets of the same size, at different correlation (co-expression) cut-offs. **D**, Diagram showing the hypothesized genetic architecture of healthy blood traits. At the core of the underlying molecular network is the set of Mendelian genes which cause blood disorders when mutated. Peripherally to the core lie regulatory genes which affect the phenotype through core genes. Cis and trans-eQTLs can give insights about cell-type specificity and can identify master regulators, i.e. genes that trans-regulate several core genes simultaneously. **E**, Proportion of network genes among Mendelian, GWAS or other genes with >1 edge, or average number of edges, at different correlation (co-expression) cut-offs. **F**, Example of a sub-network containing three Mendelian genes involved in platelets (*GP9, ITGA2B, GP1BB*). Blue genes were identified by GWAS with VEP worst consequence annotation, red genes were found in GWAS and previously known Mendelian genes, grey genes are other co-expressed genes.

Biological networks are organized hierarchically (Carlson et al., 2006; Ravasz and Barabási, 2003; Ravasz et al., 2002). The recently proposed ‘omnigenic’ model (Boyle et al., 2017; Liu et al., 2019) postulates that a small number of genes at the center (or ‘core’) of the network are more likely to be directly implicated in diseases or other phenotypes of interest, but the variants in these genes contribute only a small proportion of the overall trait heritability. Most of the trait heritability is attributable to a much larger number of ‘peripheral’ gene variants with small effect sizes that contribute to subtler physiological perturbations of phenotypes through trans-regulatory effects on core genes. It is proposed that these trans-effects can be tissue-specific or more broadly active in non-trait relevant tissues. Under the assumptions of this model, we would expect causative genes for Mendelian blood disorders to be strongly enriched at the center of the network, since they have a major impact on hematopoiesis, and trans-regulators of these genes to be enriched in the periphery (**Figure 2D**). To test this hypothesis, we accessed a manually curated list of genes causative for stem cell and myeloid disorders (SMDs, 206 genes), bleeding, thrombotic and platelet disorders (BPD, 104 genes) and bone-marrow failure syndromes (BMF 80 genes; **Supplementary Table 7**) (The NIHR BioResource, on behalf of the 100,000 Genomes Project, 2019). Mendelian (i.e. core) genes were strongly enriched in the network (149 out of 314 genes, fold enrichment (FE)=3.86, p<10^−4^, cut-off=0.7; **Figure 2C**). Furthermore, GWAS associations were strongly enriched in and near known Mendelian genes (by 2.1-fold, p=1.9 ×10^−22^), with 456 variants assigned to a Mendelian gene by VEP worst consequence annotation. Missense variants in Mendelian genes (including previously unreported variants) showed significantly greater (by 2.4-fold, p=0.02) absolute effect sizes compared to missense variants of comparable minor allele frequency (MAF) assigned to other genes (**Supplementary Figure 1C**). Moreover, Mendelian genes had more connections in the co-expression network compared to other genes, consistent with a centrality scenario (valid for co-expression cut-offs at 0.4-0.8, p ranging from 4×10^−4^to 0.02, Wilcoxon test; **Figure 2E**). Finally, the expression of Mendelian genes was significantly more correlated with other Mendelian genes (median co-expression coefficient=0.11), than random sets of genes (median=0.095, p=0.007 permutation test).

The model also predicts that common variants explain a large proportion of trait heritability through trans-regulation of core genes (Liu et al., 2019). To test this hypothesis, we accessed a large set of recently reported blood trans-eQTLs (Võsa et al., 2018). Mendelian genes were strongly enriched as targets of trans-eQTLs, compared to other GWAS genes (3.76-fold, Fisher’s p=2×10^−16^). Furthermore, first- and second-degree co-expression network neighbors of Mendelian genes were enriched for GWAS associations, representing additional potential trans-regulators -or undiscovered ‘core’-genes (p<1×10^−3^, permutation test). For example, at a correlation cut-off of 0.8, a co-expression subnetwork of 26 GWAS-associated genes was centered on three known Mendelian genes causative for spherocytosis (*SLC4A1, EPB42*) and congenital anemia (*KLF1*; **Supplementary Figure 2A**). Interestingly, these factors all play key roles in red cell cytoskeleton formation, a process regulated by *KLF1* (*Ludwig et al*., *2019*). Another example is a subnetwork containing known platelet specific genes *GP9, GP1BB* and *ITGA2B*, and eight other strongly co-expressed genes (**Figure 2F**). All of these genes are trans-regulated by the *ARHGEF3* gene, which is a known master regulator of megakaryopoiesis (Serbanovic-Canic et al., 2011).

### Blood cell trait variants map to lineage-specific hematopoietic chromatin landscapes

We next sought to delineate relevant cell states impacted by core and peripheral gene networks. To this end, we integrated all fine-mapped variants (PP_FM_>0.1%) with chromatin accessibility profiles (ATAC-seq) of 18 human hematopoietic progenitor populations (Ulirsch et al., 2019). First, we noted that fine-mapped variants falling within hematopoietic open chromatin were strongly enriched in gene targets (assigned by VEP worst consequence) compared to non-accessible variants (OR = 1.4, Fisher’s p < 2.2×10^−16^), consistent with variants acting via trans-regulation of genes in hematopoietic cell states. Next, we used g-chromVAR (Ulirsch et al., 2019), a high-resolution cell type enrichment method, to determine the hematopoietic populations most enriched for chromatin accessibility containing fine-mapped variants for 22 blood cell traits, including 6 new traits compared to a previous study in a smaller subset of the UK Biobank (Ulirsch et al., 2019). There were 43 lineage-specific enrichments surpassing experiment-wide significance (corrected for 18 cell types * 22 traits, p < 1.26 × 10^−4^) (**Figure 3A**), of which 20 were new, including novel enrichments in granulocyte-monocyte progenitor (GMP) cell subsets for variants regulating monocyte, eosinophil, and neutrophil counts.

**Figure 3.**
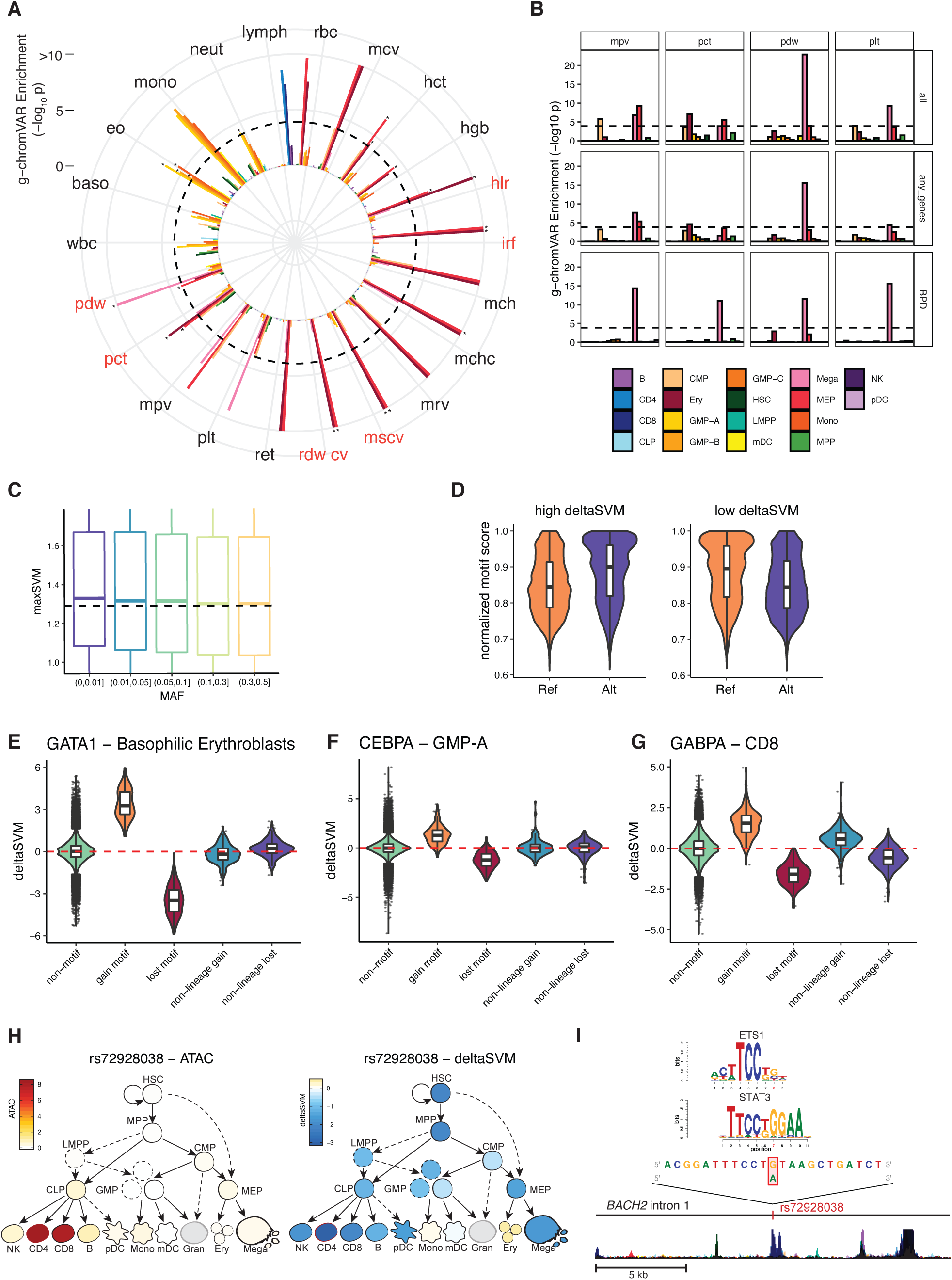
Functional annotation of fine-mapped blood trait variants. **A**, g-chromVAR results for fine-mapped variants (PP_FM_ > 0.1%) across 22 hematological traits. The Bonferroni-adjusted significance level (corrected for 22 traits and 18 cell types) is indicated by the dotted line. New traits are labeled in red. Novel enrichments are starred. The color legend for cell types is shared by both panel **A** and **B**. mono, monocyte; gran, granulocyte; ery, erythroid; mega, megakaryocyte; CD4, CD4+ T cell; CD8, CD8+ T cell; B, B cell; NK, natural killer cell; mDC, myeloid dendritic cell; pDC, plasmacytoid dendritic cell; MPP, multipotent progenitor; LMPP, lymphoid-primed multipotent progenitor; CMP, common myeloid progenitor; CLP, common lymphoid progenitor; GMP, granulocyte–macrophage progenitor; MEP, megakaryocyte–erythroid progenitor. **B**, g-chromVAR enrichment results across 4 platelet traits (mpv, mean platelet volume; pct, platelet crit; pdf, platelet distribution width; plt, platelet count), using either all trait-associated variants (all), only variants with a gene assignment (to any gene), or just variants assigned to genes causative for bleeding and platelet disorders (BPD, NIHR-RD). The original Bonferroni-adjusted significance level (corrected for all 22 hematopoietic traits and 18 cell types) is indicated by the dotted line. **C**, Association between variant absolute deltaSVM score (maxSVM), reflecting a variant’s predicted disruption of chromatin accessibility, and bins of minor allele frequency (MAF). Dotted line indicates the median maxSVM score for the MAF 0.3-0.5 bin. **D**, The allelic effects of blood trait variants with 1) high (>99th percentile) vs. low (<1st percentile) deltaSVM scores and 2) one or more predicted motif disruptions, on normalized motif scores. The normalized motif score represents the score for a variant-containing sequence as a percentage of the best score that motif could achieve on an ideal sequence. **E-G**, Cell-type specific deltaSVM scores for variants disrupting the (E) GATA1, (F) CEBPA, or (G) GABPA motif compared to scores in non-motif disrupting controls and non-lineage specific cell types. Non-motif group indicates all other variants that do not disrupt the target TF. Gain or lost motif group contains variants predicted to create or disrupted the target TF motif, respectively, with the deltaSVM score for a lineage-specific cell type (erythroblast for GATA1, GMP for CEBPA, CD8 for GABPA). Non-lineage gain/lost indicates variants predicted to create or disrupt the target TF motif, but with the deltaSVM score for non-lineage-specific populations (CD8, CD4, and B cells for GATA1 and CEBPA; erythroblast and megakaryocytes for GABPA). **H**, Lymphocyte count-associated variant rs72928038 has high chromatin accessibility (left) and deltaSVM score (right) in CD4 and CD8 populations. **I**, rs72928038 is located within intron 1 of *BACH2*, and its minor allele A is predicted to break the motifs of transcription factors ETS1 and STAT3.

We then tested whether cell type enrichments for blood traits were strengthened near core genes. Indeed, when we took the subset of g-chromVAR analysis of the variants assigned to genes causative for specific blood diseases, trait-cell type enrichments increased in lineage-specificity. For example, compared to using all trait-associated variants or variants assigned to any gene, platelet traits become more specifically enriched for megakaryocytes when looking only at variants assigned to genes causative for BPD (**Figure 3B**). This suggests that in addition to their roles in Mendelian disease, core genes are also enriched for trans-regulatory variants acting on relevant traits in a highly lineage-specific manner.

Next, we sought to predict nucleotide-specific effects of variants on chromatin accessibility. To this end, we used deltaSVM, a support-vector machine classifier, to train genomic sequence features of the ATAC-seq from 18 hematopoietic cell populations (**Supplementary Figure 2B-C**), and then applied the model to predict the allele-specific, cell type-specific impact of fine-mapped GWAS variants on chromatin accessibility (Lee et al., 2015). Out of 215,694 variants with PP_FM_ > 0.001 for one or more blood traits, we identified 22,152 variants with an absolute deltaSVM score above the 99th percentile for at least one hematopoietic cell type. Variant deltaSVM score was significantly associated with MAF (**Figure 3C**), such that lower frequency variants tended to have stronger predicted impact on chromatin accessibility (linear regression p < 2.2×10^−16^). We also observed a significant positive trend between deltaSVM and fine-map PP^FM^(linear regression p = 1.0×10^−3^, **Supplementary Figure 2D**). Variants assigned to a gene by VEP worst consequence had stronger predicted effects on chromatin accessibility compared to intergenic variants (Student’s t-test, p = 1.2×10^−3^); however, there was no significant difference in deltaSVM between variants assigned to ‘core’ vs. ‘peripheral’ genes, suggesting that variant-mediated modulation of hematopoietic transcription occurs across the entire gene regulatory network, rather than disproportionately impacting core genes.

To further characterize the regulatory effects of these variants, we predicted the potential for fine-mapped variants to disrupt 426 human transcription factor (TF) motifs (Coetzee et al., 2015). Across motif-disrupting variants, alternative alleles predicted to increase chromatin accessibility (deltaSVM score > 99th percentile) had a significantly higher motif matching score compared to the reference allele (**Methods**). The reverse was also true, indicating that deltaSVM scores track with the potential for variants to break or create TF motifs (**Figure 3D**). Moreover, this trend was cell type-specific, as evidenced by the fact that variants affecting lineage-determining TFs had a higher deltaSVM score within lineage-specific cell types, such as GATA1-disrupting variants within erythroid progenitors (Wakabayashi et al., 2016), compared to other hematopoietic populations (**Figure 3E-G**). We next sought to integrate these functional annotations in order to gain novel insights into biologically relevant variants. For example, variant rs72928038 maps to intron 1 of lymphoid TF *BACH2* (Richer et al., 2016), and colocalizes with a H3K27ac histone QTL in CD4^+^T-cells (Kundu et al., 2020). Its minor allele A (MAF = 0.18) is strongly associated with decreased lymphocyte count (PP^FM^= 0.78), and it has high chromatin accessibility in the CD4^+^ and CD8^+^ T lymphoid populations. Interestingly, the minor allele has strongly negative deltaSVM scores in the lymphoid lineage and is predicted to disrupt multiple TF motifs at the *BACH2* locus, including those with known roles in lymphocyte development, such as STAT3 and ETS1 (**Figure 3H-I**). This variant has been previously implicated in risk for several autoimmune conditions including rheumatoid arthritis (McAllister et al., 2013) and vitiligo (Jin et al., 2016). These convergent lines of evidence suggest that rs72928038 may exert effects on lymphocyte counts by altering the binding of specific lymphoid TFs within T cell progenitors. Altogether, our functional characterization of non-coding blood trait variants highlights the value of incorporating lineage-specific chromatin accessibility profiles and motif disruption analyses to nominate high-confidence mechanisms at the variant level.

### Clinical impact of rare genetic variants

The large sample size and dense imputation in this study gave us unprecedented statistical power to discover variants with low MAF and to assess their impact on human disease. First, we identified 574 rare (minor allele count > 20, MAF < 1%) blood trait variants which were either conditionally independent lead variants and/or strongly fine-mapped (PP_FM_ > 0.50), of which 512 (89.2%) were previously unreported (Astle et al., 2016; Buniello et al., 2019). These variants had higher effect sizes (p<2×10^−16^, t-test) on blood traits and were enriched for protein-coding consequences compared to other variants with similar PP_FM_ and/or lead conditional independence (27.2% vs. 4.86%, χ^2^-test p < 2.2 × 10^−16^; **Figure 4A, Supplementary Figure 2E**). Remarkably, these rare variants were strongly enriched for assignment to Mendelian blood genes (OR = 3.2, Fisher’s p = 4.22 × 10^−14^), even after excluding known pathogenic variants (**Table 1**; OR = 2.9, Fisher’s p = 4.46 × 10^−11^), but were not enriched for non-Mendelian genes (OR = 1.2, Fisher’s p = 0.18). These data support the hypothesis that a small group of high-effect rare variants disproportionately affect core genes for a complex phenotype.

**Figure 4.**
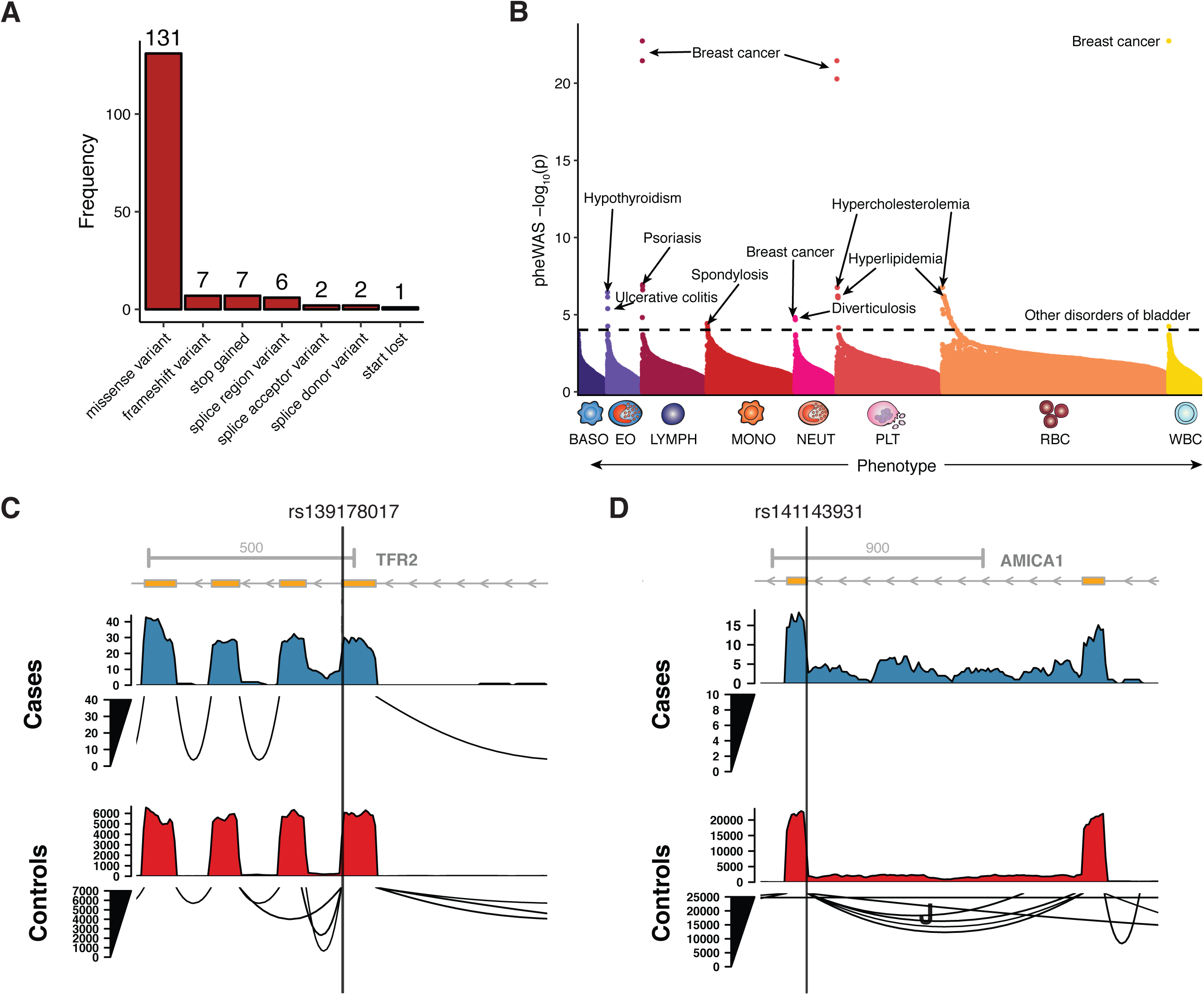
Characterization of rare blood trait variants. **A**, Distribution of coding consequences of 456 rare variants (minor allele count > 20, MAF < 1%), annotated using VEP. **B**, Phenome-wide association study of 456 rare variants (minor allele count > 20, MAF < 1%) with 529 well-represented clinical phenotypes in the UK Biobank (n up to 408,961). Variants are grouped by the hematopoietic lineage they are associated with (BASO, basophil; EO, eosinophil; LYMPH, lymphocyte; MONO, monocyte; NEUT, neutrophil; PLT, platelet; RBC, red blood cell; WBC, white blood cell). Some variants appear in more than one category if they are associated with traits from distinct lineages. The top association per category is labeled with the clinical outcome. The dotted line denotes the Bonferroni-adjusted significance level (corrected for 529 phenotypes). **C**, Sashimi plot showing donor loss splicing events in the *TFR2* locus, induced by variant rs139178017 (PP_FM_ = 0.73 for RDW, PP_FM_= 0.40 for MCV), as determined by RNA-sequencing analysis. **D**, Sashimi plot showing acceptor loss splicing events in the *JAML* (*AMICA1*) locus, induced by variant rs141143931 (PP_FM_ = 0.98 for neutrophil count).

Given the large effects of these variants on blood traits, we next sought to test their pleiotropic associations with other clinical and disease phenotypes. Thus, we performed a phenome-wide association study (PheWAS) for 456/574 rare variants using summary statistics for 529 well-represented clinical phenotypes from the UK Biobank (Zhou et al., 2018). There were 112 significant associations involving 27 variants (Bonferroni-corrected p threshold of 9.45×10^−5^; **Figure 4B, Supplementary Table 8**), of which 110 (98.2%) are currently unreported in the GWAS Catalog. Several biologically coherent associations stand out, including the missense variant rs78534766 in *ADCY7*, associated with autoimmune conditions (hypothyroidism, inflammatory bowel disease (Luo et al., 2017)) and eosinophil traits; several variants near *PIEZO1* associated with varicose veins (Fotiou et al., 2015; Van Hout et al., 2019) and erythroid traits; and a variant (rs45611741) in the 5’ UTR of *APOA5* associated with hypercholesterolemia (Nielsen et al., 2019) and MCV, and MCHC. Altogether, the PheWAS analysis revealed a variety of novel and relevant disease associations for rare blood trait variants and could point towards common mechanistic roles for these pleiotropic loci.

Splice-altering genetic variants are a prevalent and under-recognized class of variation underlying genetic disorders and complex trait regulation (Park et al., 2018). We hypothesised that a subset of variants associated with blood cell phenotypes, and particularly rare, high-effect variants, may be mediated by splicing defects. We therefore utilized the state-of-the-art neural net classifier SpliceAI to predict fine-mapped blood trait variants with splice-altering consequences (Jaganathan et al., 2019). The delta score has been shown to closely track with validation rate of cryptic splice variants, allowing it to serve as a proxy for a variant’s splice-altering probability. Across 215,694 fine-mapped variants (PP_FM_ > 0.001), we identified 109 variants with a putative splicing consequence in 106 unique genes (delta score > 0.2). Of these 109 variants, 12 (11.0%) were rare (MAF < 0.01) and confidently fine-mapped (PP_FM_> 0.50; **Supplementary Table 9**). Strikingly 85.3% (93/109) of the variants fall in non-canonical splice sites, meaning they lie outside the essential GT and AG splice junction dinucleotides. In addition, putative splice variants had lower MAF (Mann-Whitney U p = 5.08 × 10^−8^) and higher PP_FM_(Mann-Whitney U p = 9.89 × 10^−6^) compared to other fine-mapped variants (**Supplementary Figure 2F-G**). When matching by MAF and PP_FM,_these splice variants also had higher GWAS effect size (by 1.5-fold, Mann-Whitney U p=0.007). To validate these *in silico* predictions for select rare and biologically interesting novel splice variants, we examined isoform variation in RNA-sequencing data of 465 participants from the Geuvadis project (Lappalainen et al., 2013). For example, rs139178017 (MAF = 0.0053) is strongly fine-mapped for RDW (PP_FM_= 0.73) and MCV (PP_FM_ = 0.40) and predicted to induce a donor loss splice alteration for transferrin receptor 2 (*TFR2*), a partner of the erythropoietin receptor and a known regulator of erythropoiesis (Nai et al., 2015; Nandakumar et al., 2019). Compared to non-carriers, the 4 carriers of rs139178017 harbored substantially increased transcripts with intron retention adjacent to this variant (**Figure 4C**). Furthermore, we identified rs141143931 (MAF = 0.002) to be a likely causal variant for neutrophil count (PP_FM_= 0.98) and a putative acceptor loss splice variant for junctional adhesion molecule-like (*JAML*), which has been shown to mediate neutrophil migration across tight junctions (Zen et al., 2005). As expected, rs141143931 carriers in Geuvadis displayed greater intron retention downstream of the variant compared to non-carriers (**Figure 4D**). Altogether, these findings support findings that large GWAS are well-powered to identify splice variants with large phenotypic effects (Li et al., 2016), and these splice variants represent a currently under-appreciated mechanism of trait regulation in GWAS loci.

### Contribution of polygenic variation to blood cell traits and complex human diseases

Our study has identified the largest number of variants ever associated with a single group of correlated phenotypes. While each common (defined here as MAF≥1%) variant accounts for a small effect on phenotype, their joint effect may be substantial. We used different variant selection criteria to build weighted polygenic scores (PGSs) based on the UK Biobank study, and selected the one yielding most predictive power for the 29 blood measurements in an independent cohort (INTERVAL study, **Methods, Supplementary Table 10**). Remarkably, PGS based on hundreds of common sentinel variants (135-689 variants depending on trait) were shown to be more predictive than larger SNP sets employing more liberal significance thresholds, in line with findings for autoimmune diseases (Abraham et al., 2014) but contrary to evidence for other common human diseases (Khera et al., 2019). The proportion of phenotypic variance explained (R^2^) by the PGS ranged between 2.5% for basophil counts to 27.3% for mean platelet volume. Estimates obtained for the same score in an independent cohort of 2,314 French Canadians (CARTaGENE) for 15 available traits were broadly comparable, confirming portability of the PGS between European-ancestry groups (**Figure 5A**). The causal relationship between genetic variants determining eosinophil counts and asthma risk has been previously demonstrated (Astle et al., 2016). Focusing on this exemplar disease, we can show that the eosinophil count PGS was also significantly associated with asthma incidence in UK Biobank (OR=1.17, 95% CI=1.13-1.21, p=1.02×10^−19^), suggesting the potential utility of PGSs for blood biomarkers in the clinic.

**Figure 5.**
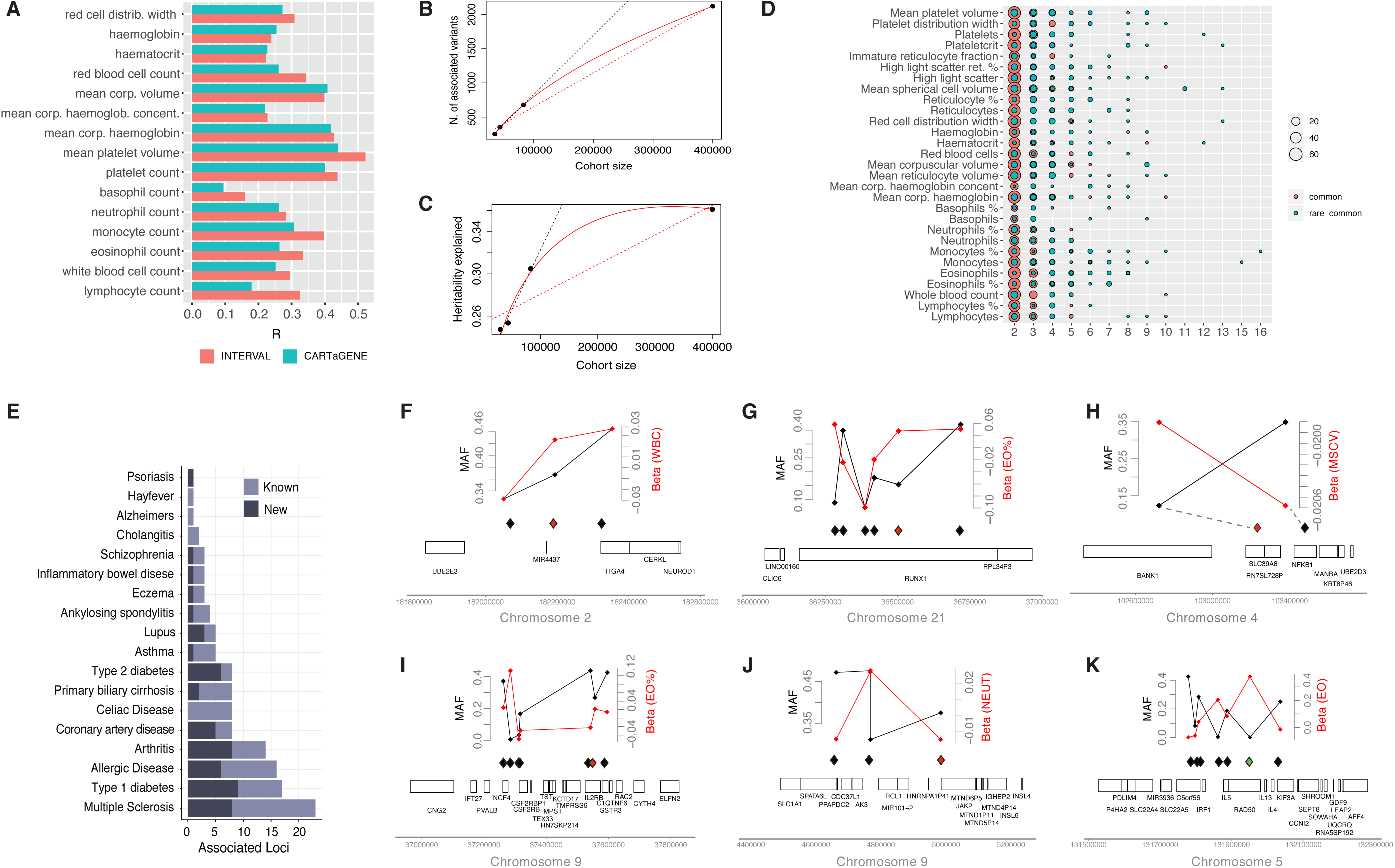
Polygenic prediction of blood traits and contribution to common diseases. **A**, Portability of the PGS across populations with European ancestry for fifteen available traits. The red bar represents the Pearson’s correlation between the score and the trait in the validation cohort (INTERVAL). Blue bars show the same in a French Canadian cohort called CARTaGENE; **B-C**, Saturation analysis showing the number of discovered variants (**B**) or proportion of heritability explained (**C**) as a function of GWAS sample size for mean platelet volume. The black dotted line is a linear projection of the first 3 points, the red dotted line is a linear interpolation of all points, the red solid curve is the best model fitting the 4 points; **D**, Loci with multiple independently associated variants for each trait. The size of the dot represents the number of loci with multiple conditionally independent variants (x-axis), the color indicates if the variants are all common (red) or a combination of common and rare (blue); **E**, Number of disease loci colocalizing (posterior probability (PP)>90%) with at least one blood count locus. Abbreviations are T2D: type 2 diabetes, PBC: primary biliary cholangitis, CAD: coronary artery disease, T1D: type-1 diabetes; **F-K**, Examples of loci with multiple conditionally independent variants associated with blood cell counts, and with at least one disease-colocalization (red diamond) or PheWAS association (green diamond) for the following genes and diseases: *ITGA4* and Inflammatory Bowel Disease - IBD (**F**), *RUNX1* and Rheumatoid Arthritis (**G**), *NFKB1* and IBD (**H**),*IL2RB* and T1D (**I**), *JAK2* and IBD (**J**), *IL4* and asthma (**K**). On the top panel, the MAF (black) and effect size (red, in SD) of each variant is plotted as a function of the variant’s position in the genomic interval.

Intriguingly, the behavior of the PGSs also suggests that the current discovery sample sizes may have achieved saturation of biological signals for blood cell traits. To begin to test this hypothesis, we modelled different discovery measures (total number of variants, loci, genes, and heritability explained) as a function of increasing discovery sample sizes. The best fitting model shows a quadratic rate of discovery decrease across all tested measures and phenotypes (**Supplementary Figure 3A-D**). However, while the total numbers of associations detected does not seem to asymptote, the heritability explained does (**Figure 5B-C**), showing that any GWAS in larger sample sizes will provide new discoveries, albeit of smaller and smaller effects, with the exception of unobserved rare variants. Greater independent discovery datasets will be required to conclusively validate this observation in the future.

Finally, we wondered if multiple conditionally independent signals at a single locus could underlie associations with complex diseases, and could help define *allelic series* at pharmacologically relevant genes. Twenty percent of blood trait loci had ≥2 conditionally independent variants, including some unusually large sets (**Figure 5D**). We overlapped these regions to colocalization results for 18 common human diseases (**Figure 5E**). **Figure 5F-K** shows six instances of such conditionally independent variant sets, of which three involve a known drug target. For example, the type I diabetes (T1D) locus tagged by rs5845323 on chromosome 9 contains one rare and six common variants, all associated with eosinophil percentage (**Figure 5I**). While the colocalizing T1D variant is intronic in *C1QTNF6* gene, the coding synonymous variant from the series is in *IL2RB* (interleukin 2 receptor subunit beta). It has recently been proposed that the cancer drug Aldesleukin (recombinant IL-2, which binds IL2RB) may be repurposed to treat T1D at low doses, and the drug is currently in phase II clinical trials for this therapeutic application (Todd et al., 2016) [ClinicalTrials.gov Identifier: NCT01862120]. There were three colocalizing loci between asthma and eosinophil count and/or percentage and a further three novel PheWAS associations of rare non-coding variants near known asthma genes (*GATA3, RAD50*, and *IL33*). One of the rare variants is part of a 270 kb set of conditionally independent variants on chromosome 5 associated with eosinophil counts, including another rare variant and five common signals (**Figure 5K**). The main players involved are *C5orf56* (*IRF1-AS1* or *IRF1* antisense RNA 1), *IRF1, IL5, RAD50, IL13, KIF3A* and *IL4*. Interestingly, both IL5 and IL4 are current therapeutic targets for treating a number of allergic diseases (Chang and Nadeau, 2017); (Ortega et al., 2014). Overall, this large set of conditionally independent variants informs future efforts to define allelic series to study genes of pharmacological importance (Claussnitzer et al., 2020).

### The influence of polygenic variation on blood disorders

Mendelian blood disorders display considerable heterogeneity in penetrance and expressivity. Furthermore, it is recognized that estimates of effect size and penetrance of pathogenic variants tend to be inflated when ascertained from patient populations (Wright et al., 2019). As shown above, the PGS defined by the common genetic variants discovered in this study explain a substantial proportion of variance of their respective phenotypes, and of common diseases as in the example of asthma. The extent to which polygenic variation contributes to the phenotypic manifestation of rare diseases remains to be determined.

To address this question, we first explored the genetic landscape of classical blood disorders in UK Biobank. We annotated each protein-coding sentinel variant using (i) ClinVar (Landrum et al., 2014), (ii) the Human Gene Mutation Database (Stenson et al., 2017) (HGMD; version pro 2018.4) and (iii) a recently curated list of variants for rare blood disorders from the Rare Disease Pilot for the 100,000 Genomes Project (NIHR-RD) (The NIHR BioResource, on behalf of the 100,000 Genomes Project, 2019). Overall, 101 sentinel variants were included in one or more databases above, of which 80% were coding, 10% were annotated to 3’ or 5’ UTRs, and the remaining were splice or intronic variants. 16 (16%) of the 101 variants were annotated to be pathogenic in either ClinVar or HGMD using strict criteria (ClinVar pathogenic with at least 2 stars or HGMD disease mutation “DM” with rank score greater than 0.1), involving 11 genes (*PKLR, HFE, HBB, PIEZO1, TMPRSS6, JAK2, MPO, CSF3R, MPL, GP9*, and *WAS*; **Table 1**). Only 5 of the 16 variants satisfied the pathogenicity criteria in both ClinVar and HGMD. Of these five, two variants previously reported as pathogenic for autosomal recessive diseases (rs116100695 in *PKLR* for pyruvate kinase deficiency of red cells and rs1800730 in *HFE* for hemochromatosis) were found in apparently healthy homozygous UK Biobank participants. Similarly we found apparently healthy homozygous carriers for another four recessive variants, reported as pathogenic in HGMD, but not in ClinVar (rs138156467 in *CSF3R* for neutropenia, rs61745086 in *PIEZO1* for dehydrated stomatocytosis and rs41316003 in *JAK2* for erythrocytosis and thrombocytosis). This lack of disease phenotype may be indicative of low penetrance, missing health record data, misannotation of these pathogenic variants, or undiscovered compensatory effects either by rare variants or background polygenic variation. For two additional recessive variants (rs137853120 in *TMPRSS6* for iron-refractory iron deficiency anemia and rs5030764 in *GP9* for Bernard-Soulier Syndrome) we observed no homozygous carriers, but heterozygous carriers were around 3 times more likely to have blood indices outside the normal range (hemoglobin <12 g/dl, platelet count < 150 × 10^9^/l), demonstrating previously unreported dosage-dependent effects (OR=3.25, 95% CI=1.85,5.37, p=5×10^−5^and OR=3.79, 95% CI=2.40-5.68, p=1.1×10^−9^ respectively)(**Figure 6A**,**B**). Data were inconclusive for the remaining eight variants, either because there were no homozygous carriers in UK Biobank (rs113403872 in *PKLR*, rs28928907 in *MPL*, rs33946267 in *HBB*, rs146220228 in *WAS*), or because the disease is typically associated with mild symptoms that are not easily detectable even in homozygous carriers (rs35897051 and rs119468010 in *MPO* for myeloperoxidase deficiency). With some caveats discussed later, this helps us refine estimates of penetrance of known and predicted pathogenic variants in UK Biobank.

**Figure 6:**
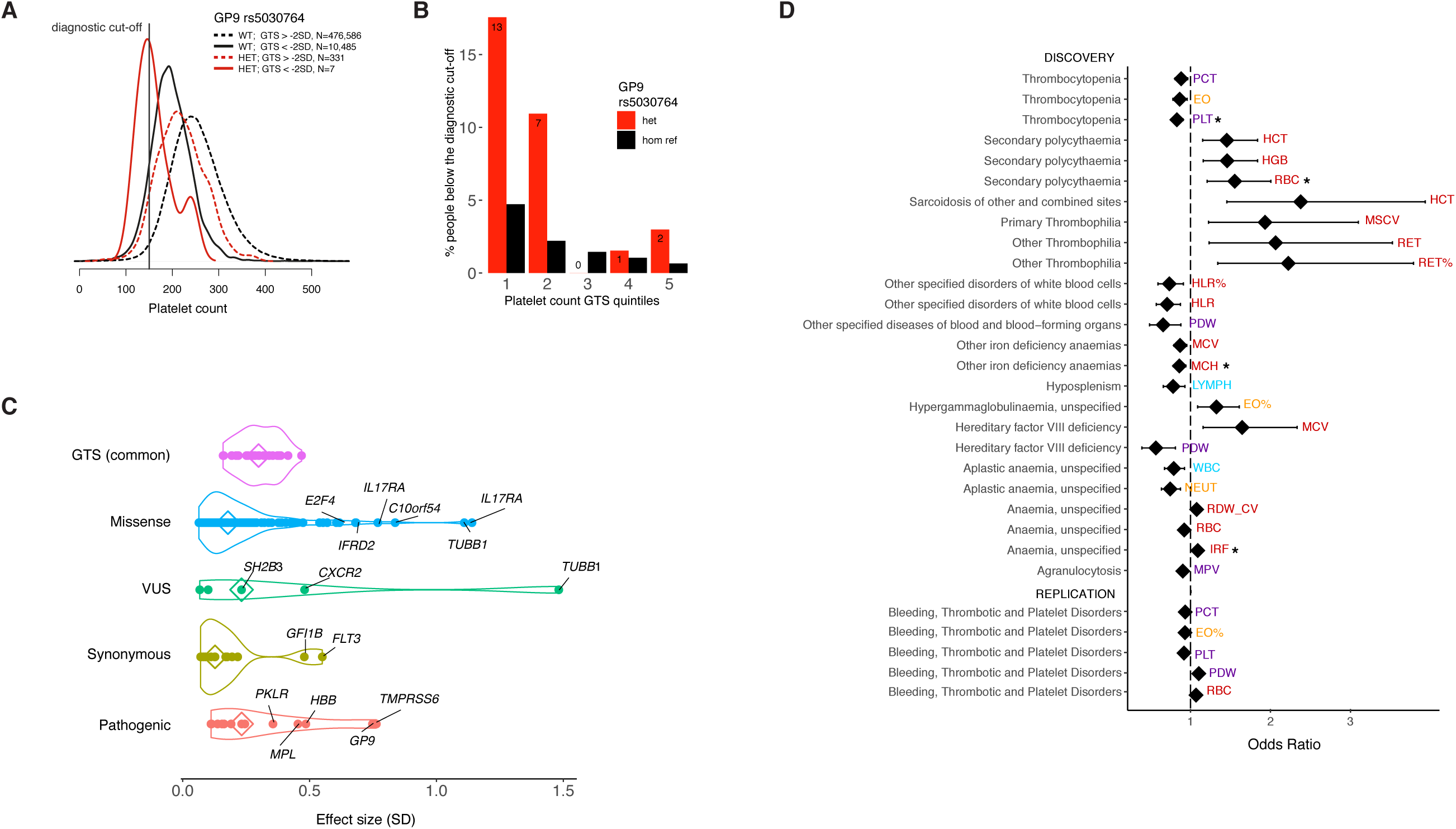
Contribution of polygenic and rare variation to rare diseases of the blood. **A**, Density distribution of platelet counts (10^9^ / liter) for UK Biobank participants who are heterozygous carriers (HET, red line) or wild-type (WT, black line) of the *GP9* rs5030764 c.182A>G (p.Asn61Ser) variant pathogenic for Bernard Soulier syndrome, plotted for participants whose PGS is above or below 2 standard deviations of the population platelet PGS. **B**, Proportion of participants below the normal range for platelet counts (150×10^9^/l) depending on PGS quintiles or *GP9* rs5030764 carriage status. **C**, Absolute effect sizes comparison between different rare variant annotations and the common polygenic score. Some of the previously unreported missenses show high effect sizes comparable to known pathogenic ones, being putative new pathogenic candidates. The contribution of the polygenic score is comparable to that of a pathogenic variant in heterozygosity; **D**, Forest plot showing the association of PGS with rare blood disorders, top 30 results are shown. Significant associations, after Bonferroni correction, are indicated by the * symbol. Diamonds represent odds ratios and whiskers show the 95% confidence interval.

We next compared the effects of PGS and rare monogenic variants. The average effect of each standard deviation of PGS ranged from 0.16 to 0.47 SD (depending on trait) and was thus comparable to that of a rare coding variant carried in heterozygosity (**Figure 6C**). We hypothesised that in the previous examples, a low penetrance of rare disease could be explained by background polygenic variation, for instance in cases where the rare disease mutation carriers have a polygenic effect in the opposite direction that compensated for a high impact rare mutation. However, the PGS of identified homozygous rare variant carriers was not different from the population mean (defined arbitrarily as PGS>2SD_PGS_ or tested by logistic regression for variants with more than 10 homozygotes). Hence, the polygenic effects alone were not sufficiently extreme to explain the lower disease prevalence in homozygous rare variant carriers.

Comparing effect sizes can also be used to screen for potential new pathogenic mutations. Variants of uncertain significance (VUS) and missense variants showed a broad distribution of effect sizes, with tails approaching the range of pathogenic variants, and could harbor putative new pathogenic variants. Among the 16 missense variants with the largest effect sizes, two previously uncharacterized variants were in known Mendelian genes (rs139473150 in *TUBB1*, associated with platelet counts and rs201514157 in *SPTA1* associated with immature reticulocytes). Platelet count was also associated with rs149254521 in *PEAR1*, a gene previously identified by an intronic variant in a platelet GWAS (Eicher et al., 2016). Subsequent functional studies showed that this gene is involved in platelet aggregation which is consistent with the phenotypes observed here (Eicher et al., 2016; Keramati et al., 2019). Two missense variants associated with monocyte count (rs140221307 and rs149771513) were in *IL17RA* (CHARGE Consortium Hematology Working Group, 2016; 2016), which has been implicated in monocyte homeostasis in mice with characterization of underlying molecular mechanisms (Ge et al., 2014). Similarly, *E2F4* is known to be essential in mice erythrocyte development (Humbert et al., 2000), and here its missense variant rs61735430 was strongly associated with mean reticulocyte volume. The other variants were in *TIE1* and *PLEKHO2* associated to platelets (rs140190628 and rs143331139 respectively), *IFRD2* (rs200622087) associated with reticulocytes, *TF* (rs8177318 and rs150854910) associated with mean corpuscular hemoglobin (MCH) and volume (MCV), *CXCR2* (rs61733609 and rs55799208) with white blood cells (Auer et al., 2014), *C10orf54* (rs201859625, in *VSIR* encoding V-set immunoregulatory receptor) with monocytes and *FAM46C* (rs148397151) with MCH. The high predictive value of the PGS, alongside future complete catalogs of genes from WGS discovering additional rare and private variants, will enable robust modelling and could help explain the considerable heterogeneity observed in many presumed monogenic blood disorders.

As shown earlier, polygenic contributions of many variants with small effects can yield effect sizes on our traits comparable or larger than that of known monogenic variants in heterozygosity. We therefore sought to explore whether they affect predisposition to rare diseases of the blood. In UK Biobank, we extracted ICD10 codes for a total of 29,080 patients and controls, with 423 diseases of blood from Chapter III “Diseases of the blood and blood-forming organs”. We then considered only a subset of UK Biobank study participants that were excluded from the discovery GWAS, either because they were classified as having a rare disease or for having extreme phenotypes (defined as their phenotype being >5 SD than the population mean, **Methods**). For each of these patients we estimated weighed PGS for sentinel genetic variants (as detailed earlier) for phenotypes matched to each disease (e.g. platelet counts [PLT] for platelet disorders). We then fit logistic regression models to test the associations of the PGS with rare disorders of the blood. For the first time, we show that PGSs derived for blood parameters influence risk for several rare blood disorders (**Figure 6D**). For instance, we showed that a higher PGS for red cell count was positively associated with incidence of secondary polycythemia, a disorder characterized by elevated hematocrit (OR=1.55, 95% 95% CI=1.21-2.00, p=6.5×10^−4^). A high PGS for mean corpuscular hemoglobin was protective for iron deficiency anemia (OR=0.86, 95% CI=0.79-0.94, p=9.1×10^−4^). A high PGS for neutrophil count decreased the risk of aplastic anemia (OR=0.74, 95% CI=0.64-0.87, p=2.9×10^−4^), which manifests as cytopenias due to depletion of hematopoietic stem cells and failure of blood cell production (Pascutti et al., 2016). Finally, the platelet count PGS was negatively associated with thrombocytopenia or low platelet count (OR=0.83, 95% CI=0.76-0.91, p=3.8×10^−5^). We replicated findings for platelet related PGSs in an independent cohort of 1,199 Bleeding, Thrombotic and Platelet Disorders (BPD) patients and 7,308 controls with whole-genome sequencing (The NIHR BioResource, on behalf of the 100,000 Genomes Project, 2019) (**Figure 6D**). We showed that an increased PGS for platelet counts resulted in a protective effect against BDP disorders, including thrombocytopenia (OR=0.92, 95% CI=0.86-0.98, p=0.007). These results refine our understanding of rare blood disease heterogeneity, and the contribution of the polygenic background of an individual to the manifestations of a rare disease known to be caused by high-impact pathogenic variants.

## Discussion

Hematopoiesis is a highly regulated process intrinsic to hematopoietic stem/ progenitor cells and intermediate cell states. Genetically controlled blood cell counts and their properties serve as a valuable paradigm for studying complex trait genetic architecture (Bao et al., 2019; Tardaguila and Soranzo, 2019). Here we present the largest set of GWAS association results ever reported for a set of biologically relevant traits (16,900 associations). The magnitude of this discovery set allowed us unprecedented statistical power to explore current paradigms in complex trait genetics, as well as build a solid bridge between GWAS in general population cohorts and existing knowledge of monogenic blood disorders, largely derived from patient cohorts.

While an omnigenic model has recently been proposed to explain complex disease/ trait architecture (Boyle et al., 2017), this framework has been met with some skepticism (Wray et al., 2018). Here, we carried out a first empirical assessment of this model in the context of blood cell trait variation. We defined core genes as a set of known Mendelian genes, which have been recently curated in a large sequencing effort by the NIHR-RD study (The NIHR BioResource, on behalf of the 100,000 Genomes Project, 2019). We then used a robust network of co-expression in whole blood to show how these genes exhibit hub characteristics when compared to other sets of genes, i.e. they have a stronger and larger set of connections among themselves and to other genes. Furthermore, they significantly overlap with the GWAS genes, which in turn comprise a large proportion of the network. This emerging picture of underlying network connectivity regulating blood traits harbours potential for discovering new pathogenic genes and drug targets. As an example, we identified a subset of closely co-expressed genes (including three known platelet genes *GP9, ITGA2B*, and *GP1BB*) that includes eight potential new players and is co-regulated by the same trans-acting eQTL in the *ARHGEF3* gene. The use of a large dataset with concurrent genetic and gene expression data in cell-defined contexts will be necessary to enable a more quantitatively accurate validation of the model (Liu et al., 2019).

By integrating the identified genetic associations with large sets of functional genomic data, including eQTL and chromatin accessibility data, we refine our knowledge of how human variation impacts hematopoiesis and the developmental stages during which this variation is likely to act. We demonstrate that the incorporation of cell type-specific chromatin accessibility landscapes with transcription factor motif analyses can reveal potential variant-to-function mechanisms. In addition, we identify and highlight the largely unexplored role of splice-altering variants as a mechanism of blood trait regulation, and validate several biologically relevant examples of associations with allele-specific isoforms through the use of transcriptomic data.

Through available large population datasets, it has become clear that rare variants can have large phenotypic effect sizes. We identified 501 rare variants, which tended to have larger effect sizes on blood cell traits and were enriched for coding variants in genes for Mendelian blood and bone marrow disorders, even after excluding known pathogenic variants. We leveraged the power of our study to test for associations of rare variants with other complex common diseases and reported 112 associations including with asthma, IBD, varicose veins, and hypercholesterolemia.

It has become clear that polygenic variation has a substantial contribution to variation in complex quantitative traits and disease risks, sometimes yielding effects comparable to those of rare pathogenic variants. Using only the sentinel signals from our discovery GWAS, we were able to build a PGS explaining up to 28% of phenotypic variance, which outperformed a more liberal approach. This suggests that our sentinel variants approach saturation for heritability explained by common variants, which we demonstrate for several blood cell traits. We therefore explored the polygenic effects jointly with pathogenic variants and as phenotype modulators in patients with rare blood disorders. While we found 16 known monogenic variants were each associated with quantitative blood phenotypes, 52 participants homozygous for five rare recessive pathogenic variants appeared to be healthy with normal blood count and indices. This suggests that the penetrance of pathogenic variants tends to be overestimated, as recently shown (Oetjens et al., 2019). Accounting for the PGS could not explain the reduced penetrance, but our analysis may be limited by the diseases we had adequate statistical power to assess. Conversely, we observed strong allele dosage-dependent effect sizes for two heterozygous variants (previously reported as recessive), that could lead to disease especially if co-inherited with an adverse PGS. For example, heterozygous carriers of the *GP9* variant rs5030764 were three times more likely to have platelet counts below the normal range (<150K/ul), compared to homozygous reference participants. Finally, we observed a significant association between phenotype-relevant PGSs and rare blood disorders for thrombocytopenia, secondary polycythemia, anemia, and aplastic anemia, regardless of the presence or absence of known rare variants in patients. This highlights a significant modulating effect of polygenic effects on presumably monogenic disorders and lays the groundwork for future studies that aim to define the impact of genetic background on variable penetrance and expressivity observed in these blood disorders.

In summary, through the largest study of blood cell trait variation to date, we provide important new insights into how variation in regulation of blood cell parameters occurs and how such variation may contribute to the variable phenotypes observed in rare blood disorders that are presumed to have a monogenic etiology. Our findings provide a novel framework for considering an individual’s genetic background and how this may impact on the presentation of blood diseases. Finally, while our study has focused on the insights we can gain into variation in hematopoietic regulation in health and disease, it is likely that the lessons learned here will be more broadly applicable to a wide range of human diseases and traits.

### STAR Methods

## Study participants

Following the success of the Blood Cell Consortium (Eicher et al., 2016, Chami et al., 2016, and Tajuddin et al., 2016), the Blood Cell Consortium Phase 2 (BCX2) continues to identify novel common and rare variants associated with blood cell traits using imputed genotype data based on Haplotype Reference Consortium (HRC) or the 1000 Genomes Project (Phase 3, version 5) for European ancestry cohorts and non-European ancestry cohorts, respectively. BCX2 is comprised of 746,667 participants from 40 discovery cohorts and five ancestries: European, African American, Hispanic, East Asian, and South Asian. BCX2 is divided into two working groups: European that consists of 563,946 participants from 26 cohorts and that is the focus of this study, and trans-ethnic. Detailed descriptions of the participating cohorts are summarized in **Supplementary Table 2**. All participants provided written informed consent, and local research ethics committees and institutional review boards approved the individual studies.

### Genotyping, quality control and imputation

Genotyping array and pre-imputation quality control (QC) for each participating cohort is provided in **Supplementary Table 2**. Recommended genotype QC metrics included MAF (>0), call rate (>98%) and Hardy-Weinberg equilibrium p (>10^−6^). The recommended pre-imputation sample exclusion criteria (**Supplementary Table 2**) included call rate (>95%), heterozygosity rate (> median+3*IQR), gender mismatches, duplicates, and outliers from principal component analysis with reference samples from 1000 Genomes Project. All genotypes were on Genome Reference Consortium Human Build 37 (GRCh37) forward strand (http://www.well.ox.ac.uk/~wrayner/strand/). We also recommended rechecking strand and allele orientation in the variant call format files prior to imputation using checkVCF (http://genome.sph.umich.edu/wiki/CheckVCF.py). Finally we recommended using the imputation servers available at https://imputation.sanger.ac.uk/ or https://imputationserver.sph.umich.edu/ with requesting the HRCr1.1 2016 reference panel, EUR population and Quality Control & Imputation Mode. All cohorts followed the procedure for the imputation of autosomal variants except for UK Biobank (UKBB) and INTERVAL that had their genotype imputation described elsewhere (Astle et al., 2016).

### Phenotype modelling and cohort level association analyses

When possible, we excluded samples with any of the following: pregnancy (when complete blood count (CBC) done), acute medical/surgical illness (when CBC done), blood cancer, leukemia, lymphoma, chemotherapy, myelodysplastic syndrome, bone marrow transplant, congenital or hereditary anemia (e.g., hemoglobinopathy such as sickle cell anemia or thalassemia), HIV, end-stage kidney disease, dialysis, EPO treatment, splenectomy, cirrhosis and those with any of the following extreme measurements: WBC count >100*10^9^/L with >5% immature cell or blasts, WBC >200*10^9^/L, Hemoglobin >20g/dL, Hematocrit >60%, Platelet >1000*10^9^/L. For the WBC subtypes (e.g. basophils count) we used the relative count, i.e. the total WBC count multiplied by the proportion for each cell type (e.g. basophils percentage). Raw phenotypes were regressed on age, age-squared, sex, principal components and cohort specific covariates (e.g., study center, cohort, etc) if needed, WBC related traits were log10 transformed before regression modelling. Residuals from the modelling were obtained and then inverse normalized for cohort level association analysis or GWAS. All cohorts followed the same exclusions and phenotype modelling except for UKBB and INTERVAL that had their procedure described elsewhere (Astle et al., 2016). The cohort level association analyses were then conducted using a linear mixed effects model that can account for known or cryptic relatedness (e.g. BOLT-LMM (Loh et al., 2015), EPACTS https://github.com/statgen/EPACTS and rvtests (Zhan et al., 2016) with the additive genetic model.

### QC and pre-processing of cohort level association analysis results

Cohort level association analysis results went through a standard QC procedure (Winkler et al., 2014; Zhan et al., 2016) using EasyQC R package (https://www.uni-regensburg.de/medizin/epidemiologie-praeventivmedizin/genetische-epidemiologie/software/). The mapping file and allele frequency reference data (GRCh37/hg19) from HRC were used to harmonize variant names across cohorts and to check allele frequency discrepancies between cohorts and the HRC reference panel, respectively. We generated a unique ID for each variant using the form of chromosome:position_allele1_allele2 where alleles were ordered lexicographically or based on indel length as tri-allelic variants and/or indels of the same chromosome:position were observed. In addition to allele frequency plots, quantile-quantile (QQ) plots and SE-N (i.e., inverse of the median standard error versus square root of sample size) plots were also checked to detect systematic inflation, and different phenotypic variances due to mis-specified phenotype transformation or regression model, different study design or different study population, etc.

### Meta-analysis

Post-QC’ed and pre-processed European cohort results were then meta-analyzed by GWAMA (Mägi and Morris, 2010) using inverse variance weighted fixed effects approach. We applied an imputation quality filter of INFO score <= 0.4 and a minor allele count (MAC) filter of MAC <= 5 for each variant in the meta-analysis, except for three large cohorts UKBB (N = 487,409), WHI (N = 17,682) and GERA (N = 53,822), where a more stringent MAC filter of MAC <= 20 was applied to exclude extremely rare variants with extreme effects prior to meta-analysis.

### Exact conditional analysis

We performed an exact conditional analysis using a stepwise multiple linear regression (Astle et al., 2016) approach in UKBB. Stepwise multiple linear regression aims to identify the parsimonious subset of variants which explain the significant associations identified by univariate GWAS. For each blood index, the set of genome wide significant variants was partitioned into the largest number of blocks such that no pair of blocks are separated by fewer than 5Mb, and no block contains more than 2,500 variants. Blocks are generated independently for each blood cell index. For each block we identify a parsimonious list of variants explaining the signal in that block using a stepwise conditional linear regression protocol discussed below. Linear regression is performed using the fastLM from the R package RcppEigen, and run with a significance threshold of 8.31×10^−9^. The stepwise linear regression proceeds in two stages, addition and removal of variants into the model which upon convergence represents the parsimonious signal in the block. The model starts with no variants and begins as follows iterating repeatedly through the following stages:

- <u>Initialisation step:</u> From the list of all variants in the block, add the variant with the lowest P value that is also below the significance threshold (8.31×10^−9^).
- <u>Dropping:</u> Study the p-values for all variants in the model, if any of these are above the significance threshold we iteratively prune and rebuild the model starting with the variant with the highest p-value. Once a variant is pruned it is returned to the list of variants not currently in the parsimonious model and may rejoin at a later iteration.
- <u>Addition:</u> Test each variant* not currently in the block sequentially in the model, add the variant with the lowest p-value which is below the threshold. Any tested variants which have a p-value of higher than 0.01 are not tested again in future iterations. * Variants are not permitted to be tested in the model if they have a LD r^2^ score of higher than 0.9 with any variant currently in the model.
- <u>Completion:</u> If the algorithm could neither add a variant into the model nor remove a variant from the model then we abort the iteration with the model at this stage representing the parsimonious model for this block.

Following identification of conditionally significant variants in each block, all conditionally significant variants within each chromosome are put into a single linear model and tested with the same multiple stepwise linear regression algorithm as that defined above. The resultant set is the ‘conditionally significant’ list of variants for the blood cell index, also referred to as “sentinel variant” throughout the text.

### Assessment of Meta-analysis conditionally significant associations and replication

Using conditional and joint analysis as implemented in GCTA, we identified independent association results in the meta-analyses at p<5×10^−9^. To define novel associations, we tested these variants using the same exact multivariate approach as above, in the UKBB, while conditioning on the variants identified by the previous step. Finally, we checked replication in the Million Veteran Project Cohort (Gaziano et al., 2016), for chromosome 1 and 3 traits, one per each major cell type (platelet counts, lymphocyte counts and red cell counts). The replication significance threshold was set to a nominal level (p<0.05) and same direction of effect.

### Fine-mapping

Statistical fine-mapping was performed in the UKBB cohort, using FINEMAP v1.3.1 (Benner et al.) [http://www.christianbenner.com/]. Input windows were defined as +-250 kb from a conditionally independent signal. In case of multiple sentinels generating overlapping windows these were merged together, resulting in window size ranging from 500kb to 1.38Mb. The number of conditionally independent signals in each window was used as prior knowledge for the maximum number of causative variants to be searched (--n-causal-snps option) and the prior standard deviation for effect sizes was set to 0.08 (--prior-std option). The LD structure was computed from the same samples included in the GWAS analysis. 95% credible sets were defined as minimal sets of variants jointly covering at least 95% of the posterior probability of including the true causative signals.

### eQTL Colocalization

We performed colocalization using *gwas-pw* (Pickrell et al., 2016) between GWAS of 10 hematological traits and transcriptomic profiling of Platelets, CD4, CD8, CD14, CD15, and CD19 cells. Where MPV, PDW, PLT#, and PCT were colocalized with eQTLs from Platelets, NEUT# and NEUT% were colocalized with eQTLs from CD15 cells, LYMPH# and LYMPH% were colocalized with eQTLs from CD4, CD8, and CD19 cells, and MONO# and MONO% were colocalized with eQTLs from CD14 cells. Loci of colocalization were defined by the recombination regions identified by (Berisa and Pickrell, 2016). Colocalization was only performed if the locus contained a conditionally independent variant in LD r^2^ > 0.8 with the eQTL sentinel. Results were filtered to include only those with posterior probability for colocalization higher than 80% resulting in a set of colocalised loci which are considered ‘highly likely’ to be colocalized (Sun et al., 2018).

### Co-expression network

A co-expression matrix computed from whole blood of 2,168 participants was used (Nath et al., 2017). The matrix quantifies correlations between genes, replicated across 2 different cohorts. The edges between genes were defined by imposing variable hard cut-offs on co-expression coefficients, e.g. two genes are linked in the network if their co-expression is higher than the cut-off. The following cut-offs were used (0.05, 0.1, 0.2, …, 0.8) but the results did not generally depend on the specific cut-off (unless otherwise stated). The overlap enrichment between GWAS genes and network genes was computed by random permutations of gene sets. The numbers of links per gene were compared between Mendelian genes and all other genes by Wilcoxon test. For co-expression among Mendelian genes, median absolute co-expression coefficients were computed for equal sized random draws of genes. The enrichment of trans-eQTLs targeting Mendelian genes was computed by Fisher test, comparing the proportion of Mendelian genes targeted by at least one trans-eQTL to the same number for other GWAS genes. The enrichment of GWAS genes among first and second degree neighbours to Mendelian genes was computed as follows: i) determine the list of neighbouring genes based on the specific cut-off, ii) intersect with 1000 random permutations of gene sets of the same size as the GWAS list, iii) compare to the actual intersection. The second degree neighbours were defined as neighbouring genes to all first-degree neighbours.

### g-chromVAR

Bias-corrected enrichment of blood trait variants for chromatin accessibility of 18 hematopoietic populations was performed using g-chromVAR, whose methodology has been previously described in detail (Ulirsch et al., 2019). In brief, this method weights chromatin features by fine-mapped variant posterior probabilities and computes the enrichment for each cell type versus an empirical background matched for GC content and feature intensity. For chromatin feature input, we used a consensus peak set for all hematopoietic cell types with a uniform width of 500 bp centered at the summit. For variant input, we included all variants with fine-mapped PP_FM_ > 0.1%.

### DeltaSVM

DeltaSVM is a machine learning model which uses sequence composition to predict cell type-specific open chromatin. It then uses this sequence vocabulary to predict the change in chromatin accessibility from each variant. We trained on two ATAC-Seq datasets: 1) 18 hematopoietic populations sorted from bone marrow, and 2) 8 stages of primary erythroid differentiation. For each dataset, we trained on strong ATAC peaks in the >80th percentile of counts matrix from each cell type. Standard 5-fold cross-validation was used to calculate AUROC.

### Transcription factor motif analysis

Prediction of the effects of fine-mapped variants on transcription factor binding sites (TFBS) was performed by using the motifbreakR package and a collection of 426 human TFBS models (HOCOMOCO) (Kulakovskiy et al., 2018). For 115,609 fine-mapped variants with PP_FM_ > 0.1%, we applied the ‘information content’ scoring algorithm and used a p cutoff of 1 × 10^−4^ for TFBS matches; all other parameters were kept at default settings.

### Phenome-wide association study

To identify associations between blood trait variants and clinical phenotypes, we conducted a phenome-wide association study (PheWAS) using summary statistics of 1,403 clinical phenotypes analyzed from the UK Biobank (see URLs). As input, we started with 574 rare variants with 0.00005 < MAF < 0.01 which were either conditionally independent lead signals or had fine-mapped PP_FM_ > 0.50. To avoid studying phenotypes with too few cases to capture these low allele frequencies, we only included phenotype-variant results for which the expected_case_minor_AC, calculated as 2 * variant_MAF * num_cases, was greater than 25. This resulted in the final inclusion of 529 clinical phenotypes (case numbers ranging from 1,236 - 77,977) across 456 variants which had pheWAS data. The Bonferroni-corrected significance threshold for pheWAS was calculated as 0.05 / 529 phenotypes = 9.45×10^−5^.

### Splice variant analysis

To predict splice variants, we used SpliceAI, a deep neural network that accurately predicts splice junctions from genomic sequence (Jaganathan et al., 2019). We obtained prediction scores for all possible single nucleotide variants in the reference genome, which were released along with the SpliceAI tool, and extracted scores for all variants with fine-mapped PP_FM_ > 0.001 in one or more blood traits from the UK Biobank GWAS. We considered a variant to have a putative splicing consequence if it had a delta score > 0.2 for one or more splicing consequences (acceptor gain, acceptor loss, donor gain, donor loss); this threshold was shown to be enriched for splice variants and have high sensitivity.

Validation of SpliceAI predictions was performed using RNA-seq data on lymphoblastoid cell lines (LCL) of 465 participants from the Geuvadis project 23. We aligned paired-end reads to the hg19 reference genome with STAR, allowing for novel splice junctions. Read alignments were visualized in the form of coverage tracks and sashimi plots using the GViz R package.

### Polygenic scores

The polygenic scores (PGSs) were computed as weighted sums of genotypes, weighted by their effect size on the phenotype (beta coefficient), using the PLINK score function. Beta coefficient estimates were computed in UK Biobank and PGS scores were tested in an independent cohort (INTERVAL). Positive effect alleles were included in order to get a positive contribution for each carried allele and consequently a positive correlation with the phenotype. The following SNP inclusion criteria were compared: a) all genome-wide SNPs after LD pruning with PLINK at 0.8 cut-off; b) LD-pruned SNPs with GWAS p <(0.05, 5×10^−4^, 5×10^−6^, 5×10^−8^); c) conditionally independent variants and fine-mapped variants with posterior probability >0.5; d) conditionally independent variants. Resulting PGSs were then standardized and Pearson’s R coefficients of correlation between each PGS and its relevant trait were computed for comparison. The phenotypic variance explained was computed as R2. Validation in a further independent cohort of French-Canadians (European ancestry) called CARTaGENE was performed using the same protocol, for the best performing PGS (conditionally independent variants). A linear regression model between the adjusted phenotypes and the PGS, adjusted by sex, age and principal components, was used to determine the PGS’s effect sizes per SD.

### Discovery saturation

To explore discovery saturation we chose 4 large GWAS analyses in cohorts of increasing sample size: INTERVAL (N∼35k), UK BiLEVE (N∼43,5k), UK Biobank 1st release (N∼83k) and UKBiobank full cohort (N∼400k). For all of these we had conditionally independent associations identified by the same method as described above. For each trait and cohort we determined the number of conditionally independent variants detected by GWAS; the number of genes identified by these variants (using VEP worst consequence annotation); the number of associated loci. Associated loci were defined based on LD-blocks computed in Pickerell et al. (Berisa and Pickrell, 2016) which had at least one conditionally independent signal. Firstly, a linear projection of the first 3 data points was visually inspected to determine in the 4th data point fitted the expected. Then 4 different regression models were tested to determine which one best described the full dataset: i) y∼x; ii) y∼sqrt(x); iii) y∼sqrt(x)+x; (iv) y∼log(x). Here y represents the counts (number of variants/genes/loci associated) and × represents the cohort size. Similarly we computed the heritability explained by the set of variants identified by each cohort and searched for the best fitting model. The heritability was computed as R^2^ of the multivariate model including the relevant variants in the full UK Biobank cohort.

### Allelic series and Disease Colocalization

We performed pairwise colocalization analysis between GWAS studies of 28 hematological parameters from the UK Biobank cohort and 18 different autoimmune and inflammatory related disorders. Our analysis was performed using summary statistics collected following GWAS of the respective studies. An inner merge was performed with variants tested for each hematological parameter and each respective disease risk GWAS. Colocalization analysis was then performed following the same protocol described above for eQTLs. Allelic series were defined as fine-mapping blocks including 2 or more associated sentinels.

### Polygenic contribution to rare blood disorders

We used UK Biobank participants with ICD10 codes in Chapter III (“Diseases of the blood and blood-forming organs”, D500-D899 codes), who were excluded from the GWAS discovery. After computing the PGS, we performed a logistic regression for the disease status, including sex, age, 10 principal components and any other co-occurring blood disorders as covariates. P-values were corrected by Bonferroni correction for the number of diseases (i.e. ICD10 codes) tested. We included only ICD10 codes with at least 40 cases for a total of 49 disorders.

### Pathogenic variants annotation

Conditionally independent variants for each trait, as well as fine-mapped variants with posterior probability of being causative greater than 50% were pulled together for the pathogenicity annotation. Genes were assigned to variants by VEP worst consequence [release 84]. The set of Mendelian genes was manually curated by the NIHR-RD project (The NIHR BioResource, on behalf of the 100,000 Genomes Project, 2019). We focused on three different sources of annotations: the ClinVar database [https://www.ncbi.nlm.nih.gov/clinvar/ - accessed 5/11/2018], the Human Gene Mutation Database (HGMD) [http://www.hgmd.cf.ac.uk/ac/index.php - accessed 17/01/2019] and a manually curated list of novel pathogenic variants, produced by the NIHR-RD sequencing project (The NIHR BioResource, on behalf of the 100,000 Genomes Project, 2019). Variants were matched by chromosome, position and alleles, in GRCh37. The following parameters were considered: a) ClinVar: categorical pathogenicity assignment (yes/no/unknown), the star rating (1-4, 1 being the most uncertain and 4 the most certain) and the Review Status; b) HGMD: categorial pathogenicity assignment and Rank Score indicating the pathogenicity confidence on a continuous scale from 0 to 1, 1 being certain pathogenicity assignment and 0 being very uncertain. The set of pathogenic variants was defined with high confidence, imposing pathogenicity in ClinVar with at least 2 stars or pathogenicity in HGMD (“DM”) with rank score greater than 0.1. Variants reported by NIHR-RD for the first time were assigned to the “variants of uncertain significance” category (VUS). To assess the effects of such pathogenic variants and their penetrance, two types of data were considered: full blood count diagnostic cut-offs as used in the clinics and ICD-10 codes for blood disorders (Chapter III), as recorded by UKBB. Participants with full blood counts in the normal range and no ICD-10 code were considered healthy. The joint modelling of rare variants and PGS was performed only for variants with more than 10 homozygotes, using logistic regression and relevant covariates, as above. For variants with less than 10 homozygotes, we checked for systematic PGS deviation from the population mean (defined as abs(PGS)>2*SD_PGS_).

**Supplementary Information** is available for this paper.

**Acknowledgments, author contributions and declarations** of interests are included in the

**Supplementary Information**.

## Data availability

Summary statistics for GWAS results are available to download from ftp://ftp.sanger.ac.uk/

**Table 1.**
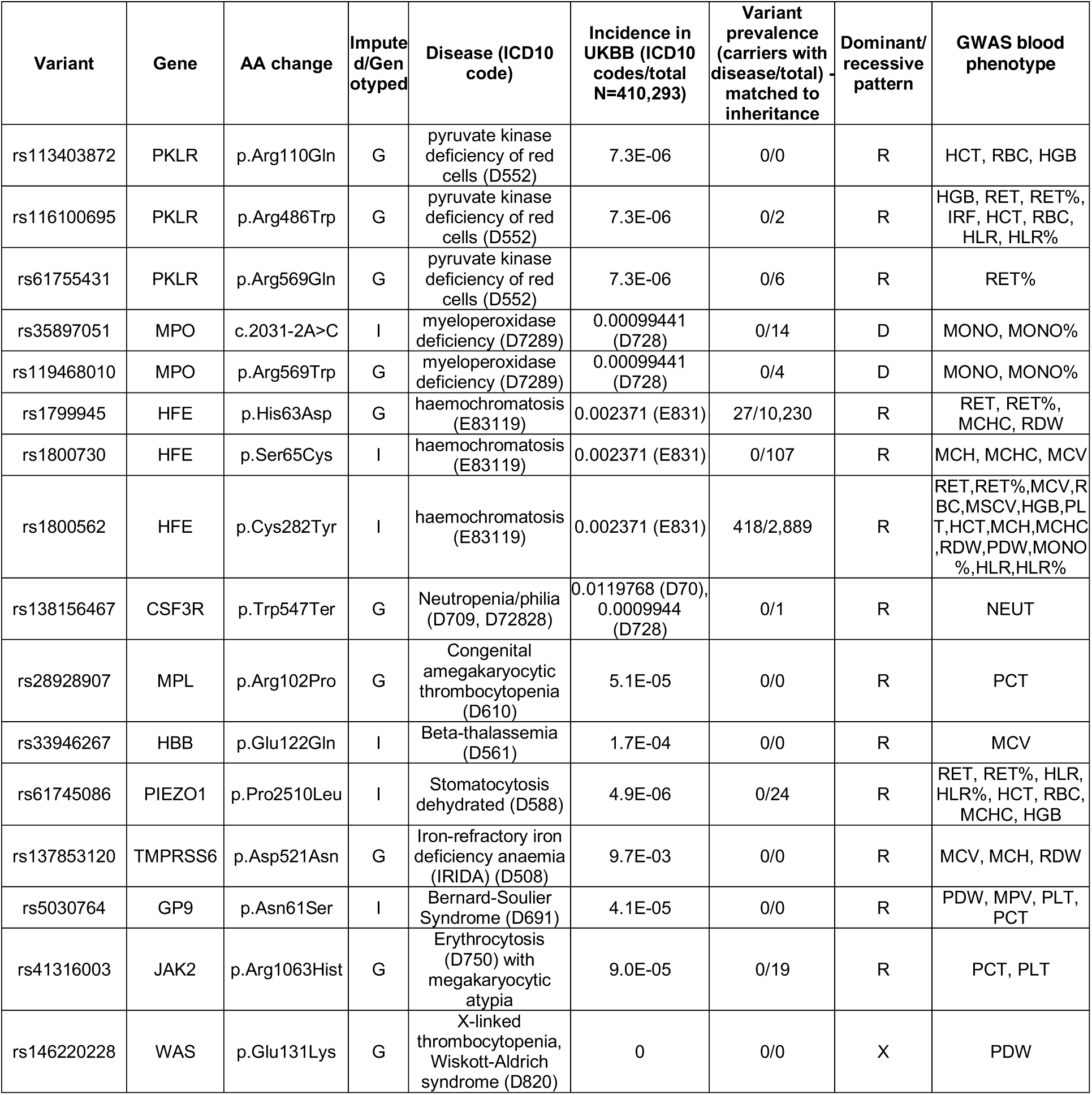
Annotation of pathogenic variants

## Supplementary Tables and Figures Legends

**Supplementary Table 1. Phenotypes’ description**

Summary of measurement methods for full blood count phenotypes used in discovery GWAS.

**Supplementary Table 2. BCX Consortium cohort information**

The study design, number of participants and sample quality control for all study cohorts is reported together with genotyping methods, SNP quality control criteria, imputation, and statistical analysis used.

**Supplementary Table 3. Conditionally independent associations in UK Biobank**

Information about each of the 16,900 variants associated and their corresponding sentinel variants is given, ordered by chromosome and position (all coordinates are with respect to GRCh37). Locus ID is a unique identifier for each locus. The column “Novel vs Astle et al. 2016” indicates if the variant or any variants in LD r^2^>0.8 was already found to be associated with the same traits in the cited previously published meta-analysis including the first release of UK Biobank. The unique variant ID is constructed from the chromosome, position and the reference and alternative alleles according to the human genome reference (build 37 coordinates). Where available, the rsID is also given. GWAS summary statistics for univariate and multivariate (conditional) model are provided, as well as the VEP worst consequence annotation.

**Supplementary Table 4. Additional conditionally independent associations from the meta-analysis**

Variants identified using GCTA-COJO that are genome-wide significant (P<5e-9) and conditionally independent from the UKBB-only association results. All joint results were calculated after regressing out the effect of the variants found in the UKBB-only analyses.

**Supplementary Table 5. Fine-mapping posterior probabilities**.

The table reports all the sentinels fine-mapped to a single causative signal, i.e. having FM_PP_> 0.95.

**Supplementary Table 6. Network connectivity by annotation**.

The co-expression network characteristics are shown based on different correlation coefficient cut-offs applied. The correlation cut-off identifies genes that are co-expressed with at least one other gene at an absolute correlation higher than the cut-off. Hence higher cut-off values identify smaller networks (columns A and B). The remaining columns show the overlap between the network and the genes identified by GWAS, based on different annotation strategies (absolute numbers and percentages are shown). Finally, column I shows how many Mendelian genes are included in the relevant network.

**Supplementary Table 7. NIHR-RD list of Mendelian genes**

The table shows a manually curated list of 314 genes known to cause blood disorders. The list was compiled by expert clinicians involved in the NIHR-RD sequencing project and is to date the most updated resource for blood disease genes and mutations. The genes are divided in 3 (possibly overlapping) categories: Stem cell and Myeloid Disorders (SMD), Bleeding, Thrombotic and Platelet Disorders (BPD) and Bone Marrow Failure (BMF).

**Supplementary Table 8. PheWAS results**.

Phenome-wide association study (pheWAS) significant associations between blood trait variants and other clinical phenotypes. var, variant ID in chromosome:position:allele1:allele2 format; ID, variant RSID; pheno, pheWAS phenotype associated with variant; trait, blood trait associated with variant; effect_allele, allele corresponding to GWAS effect sizes; blood_beta; GWAS effect size for the blood phenotype; PP_FM_, fine-mapped posterior probability; VEP_cons_most_serious, VEP most serious consequence; num_cases, number of cases of the pheWAS phenotype; maf, variant minor allele frequency; beta, GWAS effect size for the pheWAS phenotype; sebeta, GWAS standard error of the effect size for the pheWAS phenotype; pval, GWAS p-value for the pheWAS phenotype; nearest_gene, nearest gene to the variant.

**Supplementary Table 9. Splice variants associated with blood traits**. Putative splice variants fine-mapped for blood trait associations (SpliceAI delta score > 0.2 and fine-mapped PP_FM_> 0.001). var, variant ID in chromosome:position:allele1:allele2 format; rsid, variant RSID; chr, chromosome; pos, position; ref, reference allele; alt, alternate allele; SYMBOL, spliced target gene; STRAND, strand with splicing effect; DIST, distance between canonical and newly created splice sites; DS_AG, Delta score (acceptor gain); DS_AL, Delta score (acceptor loss); DS_DG, Delta score (donor gain); DS_DL, Delta score (donor loss); DP_AG, Delta score (acceptor gain); DP_AL, Delta score (acceptor loss); DP_DG, Delta score (donor gain); DP_DL, Delta score (donor loss); PP_FM_, fine-mapped posterior probability; AF_Allele2, frequency of alternate allele; trait, blood trait; frame, effect of splice variant on reading frame.

**Supplementary Table 10. PGS correlation**.

Results for all tested polygenic scores are shown as the R (Pearson correlation coefficient) between the score and the real phenotype in the INTERVAL cohort. For each set of SNPs the PGS was tested in the entire cohort and in males only (M) to ensure that no sex bias was present. Column Q shows the R^2^(i.e. the trait variance explained) for the best score, which includes conditionally independent variants only. Abbreviations: GW=genome-wide.

**Supplementary Figure 1.**
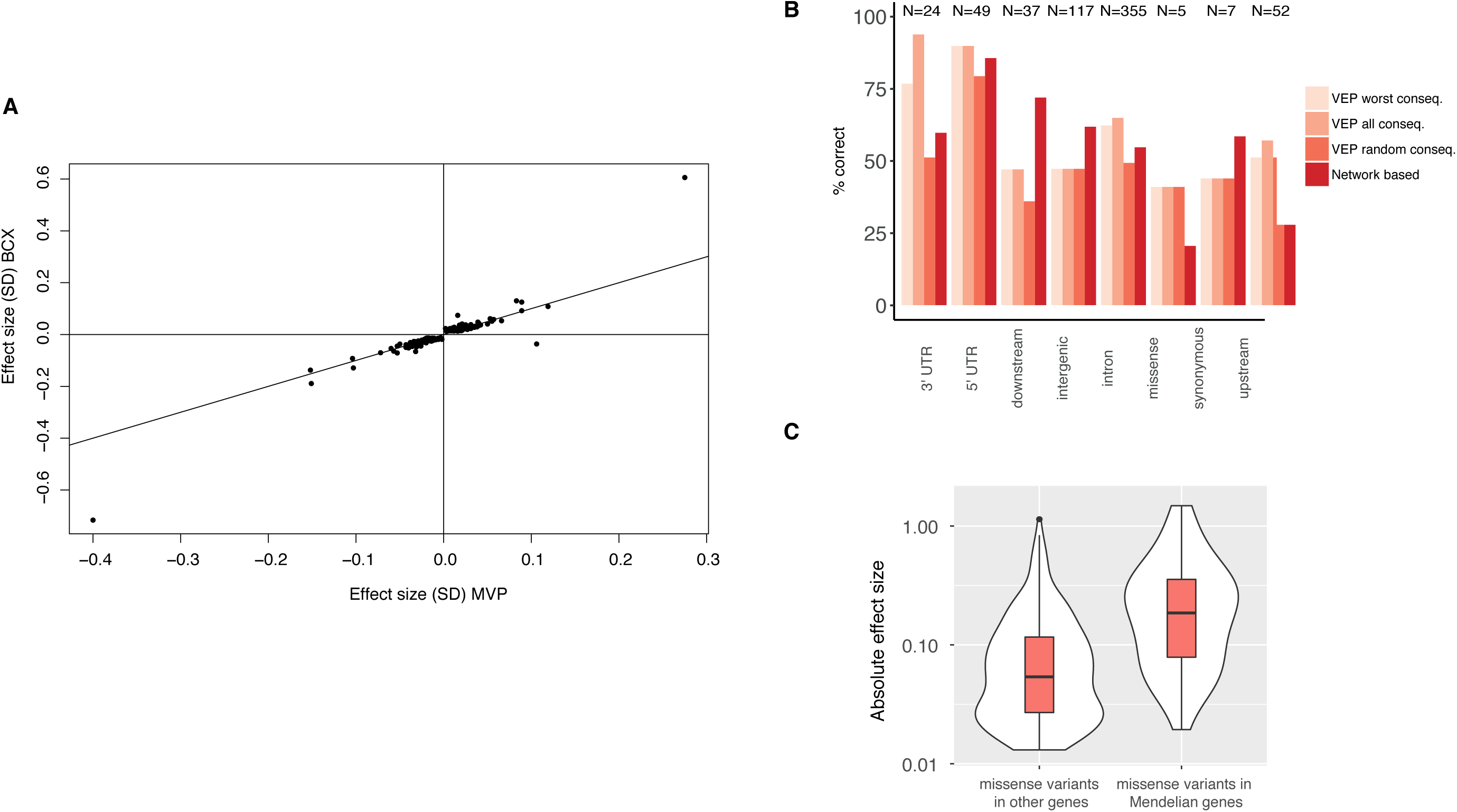
**A**, Comparison of replication effect size estimates, the y-axes shows effect sizes in UK Biobank, the x-axes shows effect sizes in MVP. **B**, Proportions of correct gene to variant assignments for four different annotation methods: VEP worst consequence and VEP all consequences, one random gene among VEP all consequences annotations and proximity-based, i.e. looking into FM-blocks (250 kb flanking sentinels) and choosing the gene that belongs to the co-expression network, if present. Only known eQTLs in matched cell-types are shown and the correct gene is assumed to be the one identified by the eQTL experiment. **C**, Missense variants assigned by VEP to Mendelian genes have higher effect sizes compared to other variants, after matching for MAF.

**Supplementary Figure 2.**
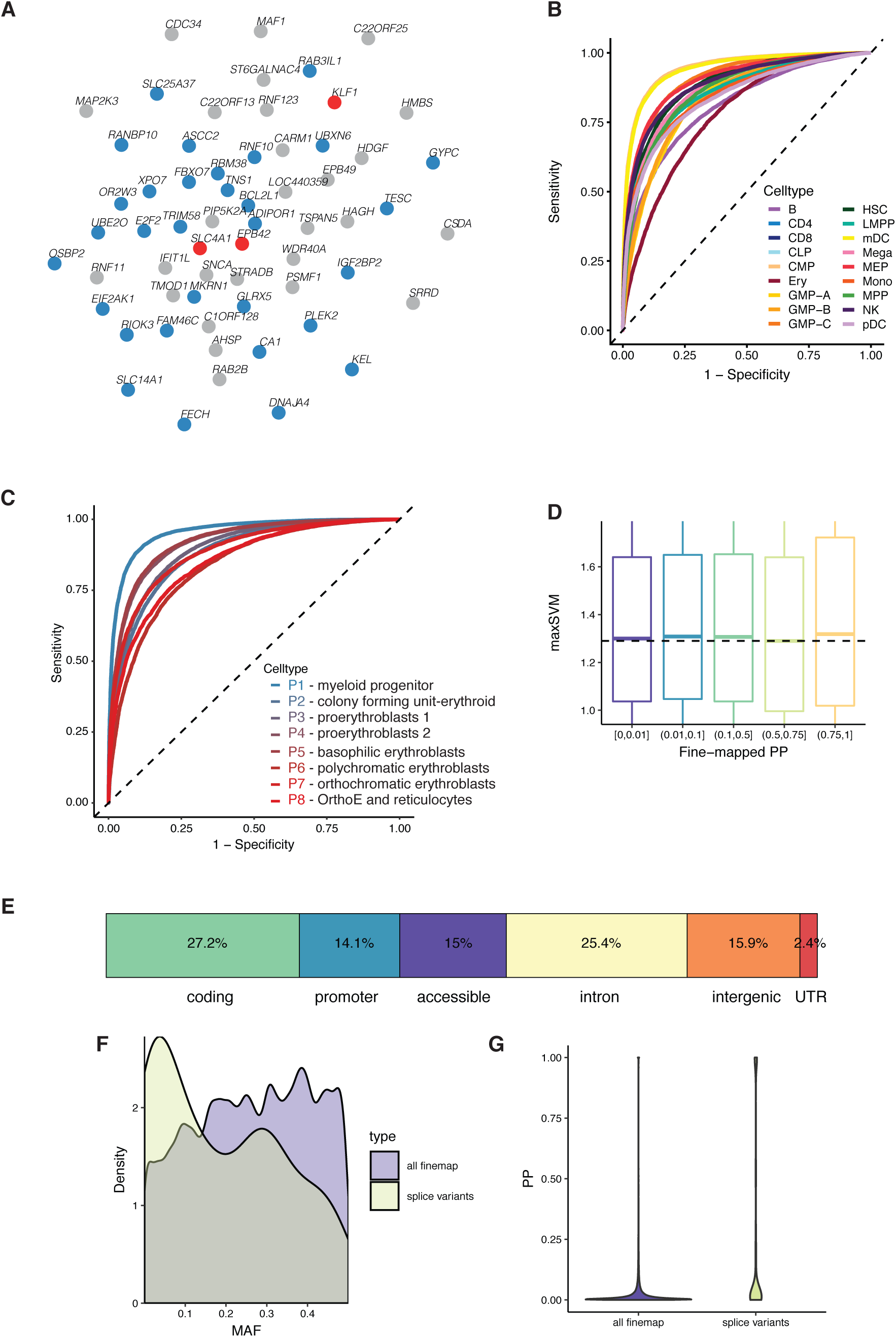
**A**, A zoom-in example of the co-expression network, including connected genes with a very high correlation cut-off (0.8). Blue dots represent genes detected by GWAS, according to VEP worst consequence annotation, red dots represent GWAS genes that are also Mendelian genes for blood disorders. Three Mendelian genes are identified, all of them involved in spherocytosis and other red-cell disorders. **B-C**, Receiver operating characteristic (ROC) curves for measuring classification performance of deltaSVM in two datasets: B) 18 hematopoietic populations sorted from bone marrow, and C) 8 stages of primary erythroid differentiation. **D**, Association between absolute variant deltaSVM score (maxSVM), reflecting a variant’s predicted disruption of chromatin accessibility, and bins of variant posterior probability (PP_FM_). **E**, Rare variants (minor allele count > 20, MAF < 1%, PP_FM_> 0.50, conditionally independent) grouped by genomic annotation. **F**, Density distribution of variant minor allele frequency (MAF), comparing 109 putative splice variants to all fine-mapped blood trait variants. **G**, Violin plot of the fine-mapped posterior probability (PP_FM_) for putative splice variants vs. all fine-mapped variants. For variants fine-mapped to multiple blood traits, we used the maximum PP_FM_.

**Supplementary Figure 3.**
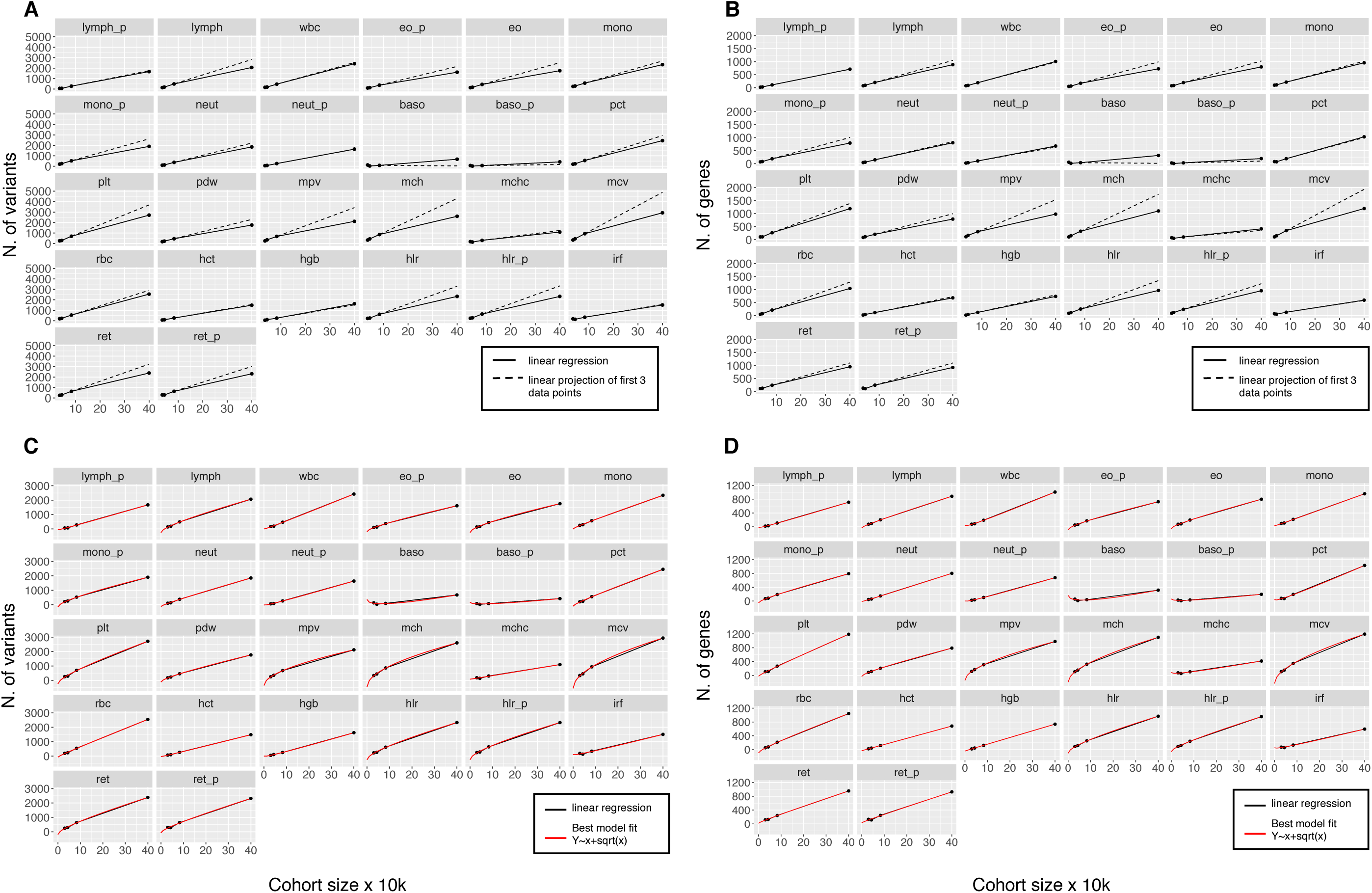
**A**, For each trait, we show the number of conditionally independent variants (y-axes) discovered by GWAS in four cohorts of increasing sample size. The sample size is shown on x-axes in 10,000s. Two linear regression lines are shown: the full black line represents a regression including all 4 data points, the dotted black line represents a linear projection of the first three data points for comparison. A decreasing trend can be observed for almost all traits. **B**, Similarly to panel (A), the number of GWAS-identified genes is shown on the y-axes. Genes were identified by VEP worst consequence annotations. **C**, The same data points as in panel (A) are now shown with the best fitting model line in red, which corresponds to a square-root growth model. **D**, The same data points as in panel (B) are now shown with the best fitting model line in red, which correspond to a square-root growth model.

## Data Availability

Summary statistics for GWAS results are available to download from ftp://ftp.sanger.ac.uk/

## Notes

### Competing Interest Statement

The authors have declared no competing interest.

### Funding Statement

Funding details are provided in the supplementary information.

